# Exploring the effects of Dasatinib, Quercetin, and Fisetin on DNA methylation clocks: a longitudinal study on senolytic interventions

**DOI:** 10.1101/2023.09.22.23295961

**Authors:** Edwin Lee, Natàlia Carreras-Gallo, Leilani Lopez, Logan Turner, Aaron Lin, Tavis L. Mendez, Hannah Went, Alan Tomusiak, Eric Verdin, Michael Corley, Lishomwa Ndhlovu, Ryan Smith, Varun B. Dwaraka

## Abstract

Senolytics, small molecules targeting cellular senescence, have emerged as potential therapeutics to enhance health span. However, their impact on epigenetic age remains unstudied. This study aimed to assess the effects of Dasatinib and Quercetin (DQ) senolytic treatment on DNA methylation (DNAm), epigenetic age, and immune cell subsets. In a Phase I pilot study, 19 participants received DQ for 6 months, with DNAm measured at baseline, 3 months, and 6 months. Significant increases in epigenetic age acceleration were observed in first-generation epigenetic clocks and mitotic clocks at 3 and 6 months, along with a notable decrease in telomere length. However, no significant differences were observed in second and third-generation clocks. Building upon these findings, a subsequent investigation evaluated the combination of DQ with Fisetin (DQF), a well-known antioxidant and antiaging senolytic molecule. After one year, 19 participants (including 10 from the initial study) received DQF for 6 months, with DNAm assessed at baseline and 6 months. Remarkably, the addition of Fisetin to the treatment resulted in non-significant increases in epigenetic age acceleration, suggesting a potential mitigating effect of Fisetin on the impact of DQ on epigenetic aging. Furthermore, our analyses unveiled notable differences in immune cell proportions between the DQ and DQF treatment groups, providing a biological basis for the divergent patterns observed in the evolution of epigenetic clocks. These findings warrant further research to validate and comprehensively understand the implications of these combined interventions.

## Introduction

Senescence is defined as a stable growth arrest of cells that can limit the proliferation of damaged cells, which is important for tissue homeostasis [1], [2]. However, senescent cells also release harmful substances that can cause inflammation and damage to nearby healthy cells [1], [3]. Recent studies suggest that senescence may contribute to aging and age-related pathologies through the impossibility of tissue renewal by the stem cells caught in senescence or through the chronic inflammation of nearby cells that can lead to tissue dysfunction [4]–[6]. In fact, studies in mice have demonstrated that injecting senescent cells can induce age-related conditions like osteoarthritis, frailty, and reduced lifespan [7], [8].

Aging is characterized by gradual functional decline [9]. It is associated with increased risk of multiple chronic diseases, geriatric syndromes, impaired physical resilience, and mortality [9]– [11]. For this reason, the pursuit of strategies to combat age-related diseases and promote healthy aging has increased in recent years.

Given the potential role of senescence in aging, senolytic drugs have emerged as promising candidates for extending lifespan. Some initially identified senolytics were Dasatinib, Quercetin, and Fisetin [12]. These molecules were drugs or natural products already used for other indications in humans, including anti-cancer therapies [13]–[15]. Dasatinib is a tyrosine kinase inhibitor approved by the FDA to treat myeloid leukemia [12], [16]. Quercetin is a flavonoid compound that induces apoptosis in senescent endothelial cells [12], [16]. Combined treatment with Dasatinib and Quercetin (DQ) has been demonstrated to decrease senescent cell burden in humans in multiple tissues [12], [17]–[20]; improve pulmonary and physical function along with survival in mice while lessening their age-dependent intervertebral disc degeneration [7], [21], [22]; and reduce senescence and inflammatory markers in non-human primates [23]. In human studies, patients with idiopathic pulmonary fibrosis, a senescence associated disease, improved 6-minute walk distance, walking speed, chair rise ability and short physical performance battery after 9 doses of oral DQ over 3 weeks [24]. Fisetin is another flavonoid compound that has gained recognition for its anti-proliferative, anti-inflammatory, and anti-metastatic properties [15], [25]. Fisetin has the potential to reduce senescence markers in multiple tissues in murine and human subjects [12], [26]. Administration of Fisetin to old mice restored tissue homeostasis, reduced age-related pathology, and extended median and maximum lifespan [26]. Notably, a comparative study has highlighted Fisetin as the safest and most potent natural senolytic among the tested compounds [26].

To date, research has not determined the effect of senolytics in biological aging measured by molecular biomarkers, such as the length of the telomeres, the proportion of immune cells, and the alteration of DNA Methylation. DNA methylation (DNAm) has emerged as a widely used biomarker for predicting health span and age-related diseases [27], [28]. The widely used first-generation clocks, such as the Hannum clock and Horvath clocks, analyze DNAm patterns to estimate an individual’s chronological age [29], [30]. Second-generation clocks, including the DNAmPhenoAge and GrimAge, instead of being trained to predict chronological age, have been trained to predict biological phenotypes, which has increased the hazard ratio prediction of age-related outcomes [31], [32]. Moreover, a third-generation clock, the DunedinPACE, measures the rate of aging rather than providing an overall age estimation [33].

Therefore, this study aims to comprehensively assess the impact of senolytic drugs on epigenetic aging through two longitudinal studies to address our research objective. The initial investigation focuses on a combination treatment of Dasatinib and Quercetin, while the subsequent phase incorporates Fisetin into the treatment regimen, allowing for the evaluation of aging effects over a two-year period. This comprehensive approach will provide insights into how senolytic drugs influence epigenetic dynamics and contribute to our understanding of potential interventions in the process of senescence.

## Results

Figure 1 shows the design of the study. 28 individuals were enrolled in our cohort and split across two different studies. In the first study, a total of 19 participants underwent a 6-month treatment period with DQ, and blood samples were taken at baseline, 3 months, and 6 months. The age range of these individuals was between 43.0 and 86.6 (**Table 1**). Following the completion of the initial treatment period, participants remained untreated for a duration of one year. Out of the initial group, 10 participants continued in the trial, while 9 new participants joined the study. In the second trial, all participants underwent a 6-month treatment with DQ and Fisetin, with measurements taken at baseline and 6 months. The average age of the participants in these two studies was 60.9, ranging from 44.5 to 88.0. The percentage of male participants varied across the studies: 57.9% in the DQ study and 42.1% in the DQF study.

**Figure 1.**
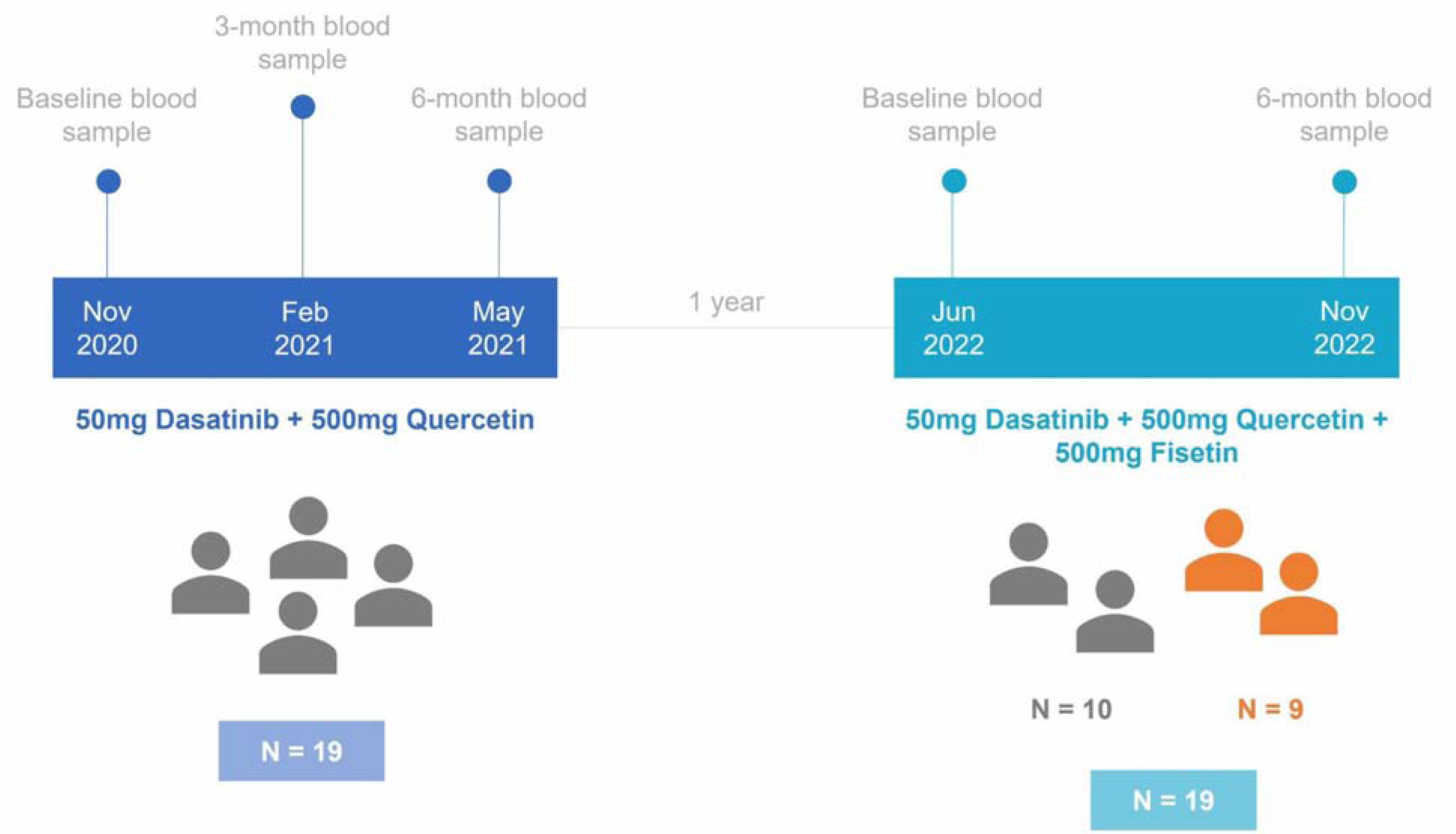
Timeline diagram for the study design. In the first study, 19 individuals were treated with 50mg of Dasatinib and 500mg of Quercetin. After one year, the second study started with 10 participants from the first study and 9 new participants. These individuals were treated with 50mg of Dasatinib, 500mg of Quercetin, and 500mg of Fisetin.

**Table 1.**
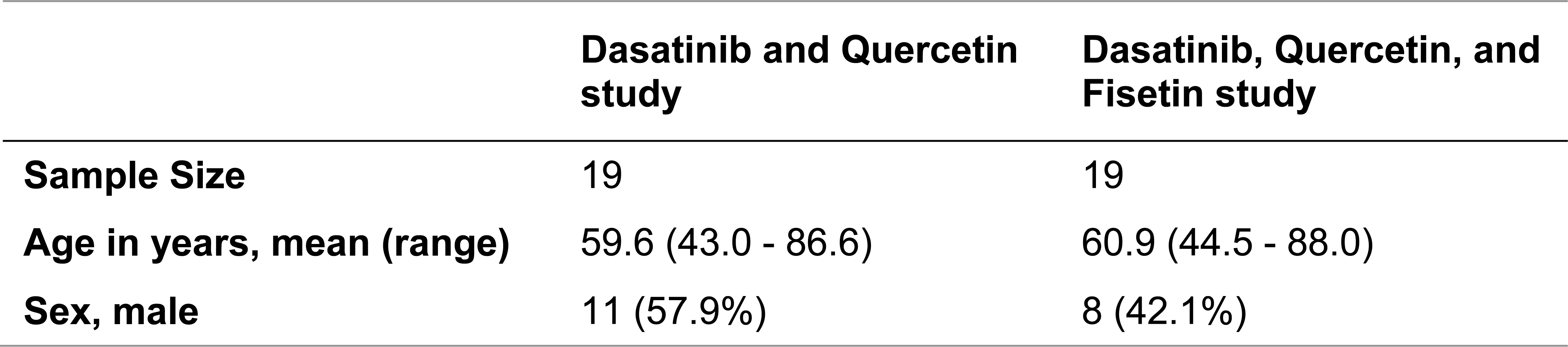
Characteristics of participants in both trials.

**Figure 2.**
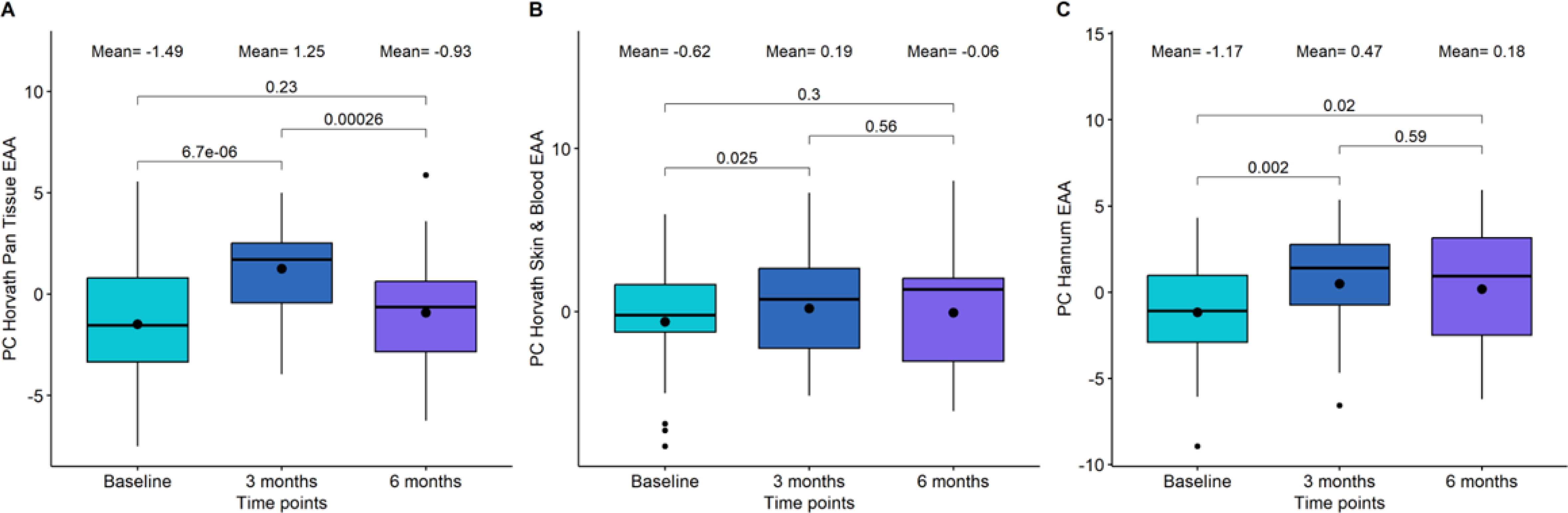
Boxplot showing the evolution of epigenetic age acceleration (EAA) first-generation clocks in the Dasatinib and Quercetin (DQ) study. (A) PC Horvath pan tissue EAA. (B) PC Horvath Skin&Blood EAA. (C) PC Hannum EAA. In the X-axis, the time points of measurements, in the Y axis, the EAA measure. On the top, the mean values at each time points. The p-values of the paired t-tests are also displayed in the plots.

**Table 2.**
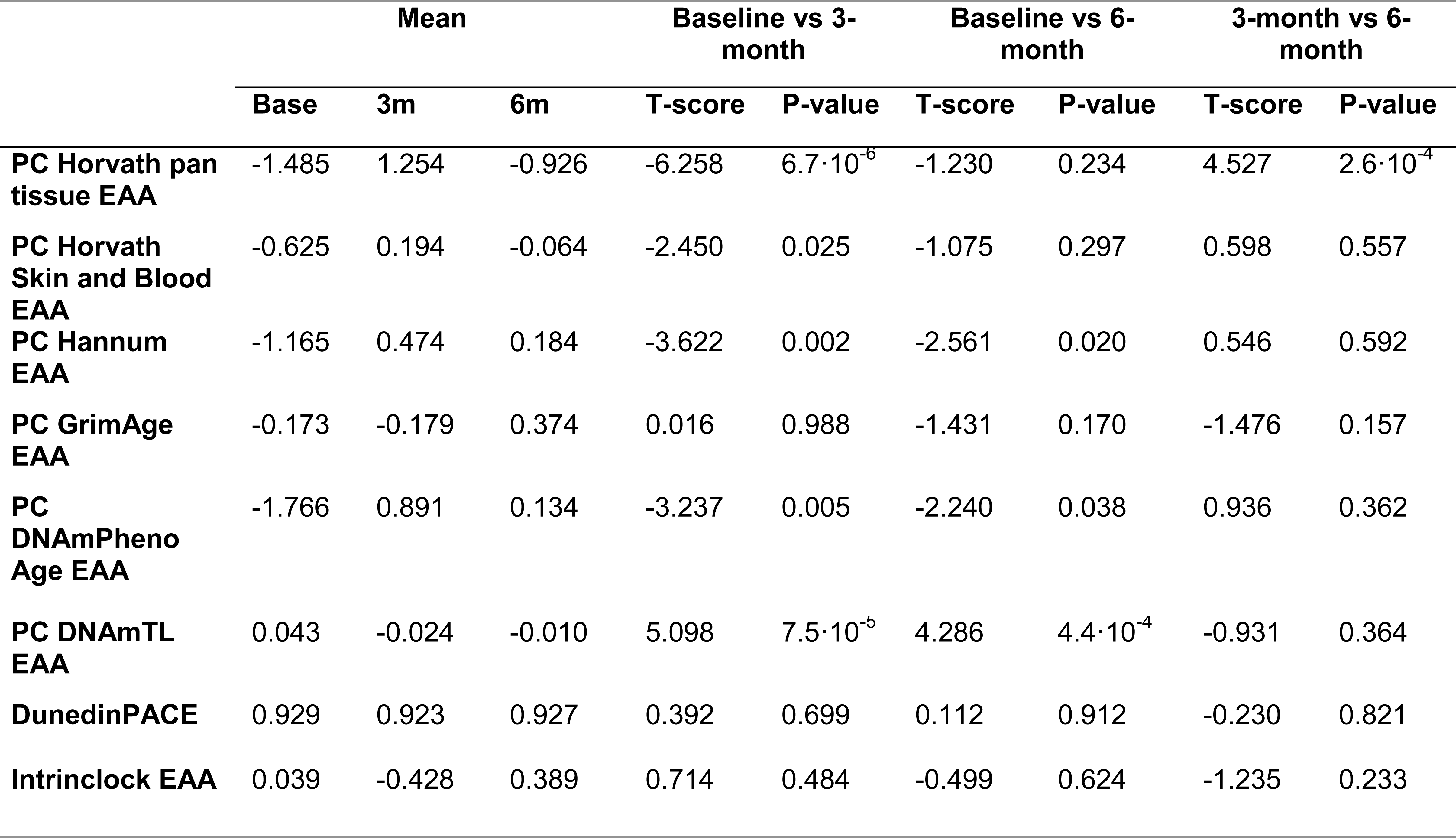
Statistical analysis for comparing baseline, 3 months, and 6 months epigenetic age acceleration (EAA) in the Dasatinib and Quercetin Study. The first three columns show the mean values for each EAA clock at each time point. The next columns have information about the t-test between baseline and 3-month test, between baseline and 6-month test, and between 3-month and 6- month tests, respectively.

**Table 3.**
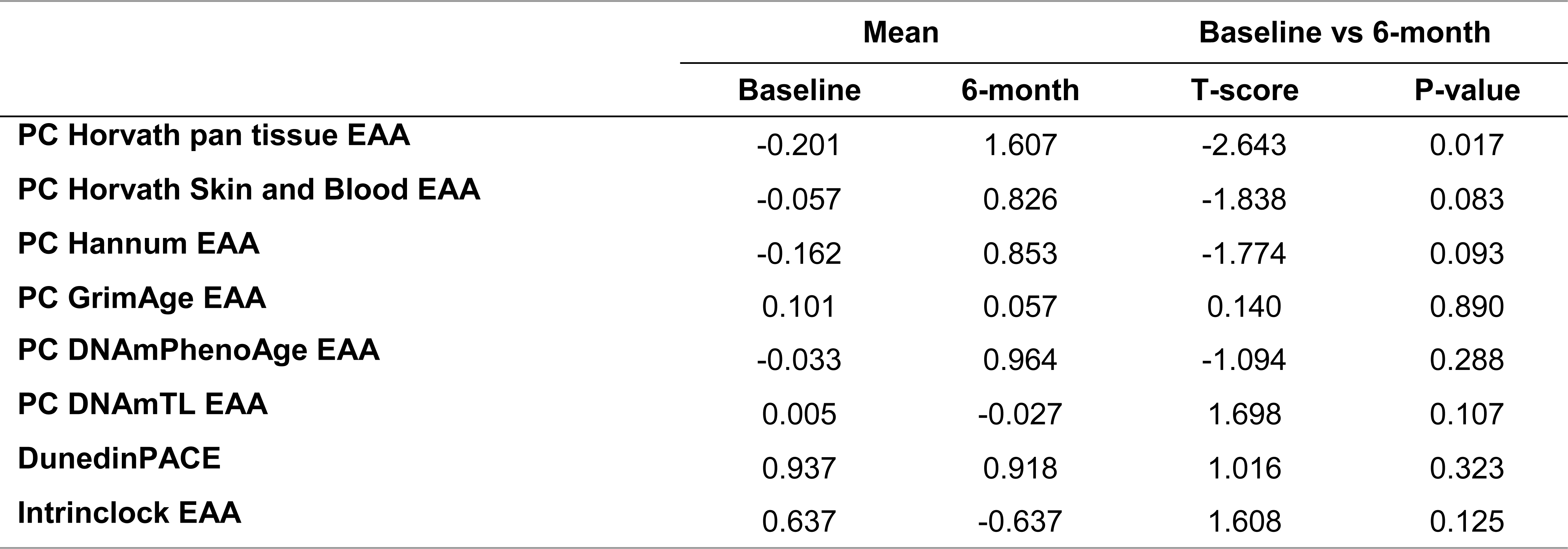
Statistical analysis for comparing baseline and 6-month epigenetic age acceleration (EAA) in the Dasatinib, Quercetin, and Fisetin Study. The first two columns show the mean values for each EAA clock at each time point. The next columns have information about the t-test between baseline and 6-month test.

### Impact of senolytic drugs on epigenetic age

To investigate the impact of the combination of DQ and the combination of DQF on epigenetic age, we assessed biological age using the principal component versions of multiple epigenetic clocks: the first-generation Horvath pan tissue clock, the Horvath skin and blood clock, and the Hannum clock; the second generation DNAmPhenoAge and GrimAge; and third generation DunedinPACE. For these clocks except DunedinPACE, we calculated the epigenetic age acceleration (EAA) after adjusting the values by age and principal components.

We observed that PC Horvath pan tissue EAA significantly increased after 3 months of DQ treatment (p-value=6.7·10^−06^). However, this increase was followed by a decrease 3 months later (p-value=2.6·10^−4^), resulting in non-significant differences between the baseline and the 6- month time point (p-value=0.23) (Figure 2A; **Table 2**). Besides, we detected a significant increase in this clock after 6 months of DQF treatment (p-value=0.017, see **Table 3**).

Regarding the other first-generation clocks, PC Horvath Skin&Blood EAA and PC Hannum EAA, we identified a significant increase following DQ treatment, particularly at the 3-month period (Figure 2B-C; **Table 2**). However, we did not observe significant changes in EAA after DQF treatment for these clocks (**Table 3**).

For second-generation clocks, we observed a significant increase between baseline and 3- month test after DQ treatment in PC DNAmPhenoAge (p-value=0.005) and between baseline and 6-month test (p-value=0.038). On the other hand, PC GrimAge and the third-generation clock, DunedinPACE, remained stable after the treatment. For DQF treatment, all the second and third-generation clocks were unchanged.

We also evaluated the changes observed by the recently developed IntrinClock, as it is agnostic to immune cell changes that have been shown to influence the reliable quantification of epigenetic age [34]. However, no significant differences were observed in any of the trials.

### Impact of senolytic drugs on DNAm Telomere length and Mitotic Clock Epigenetic Methylation Prediction Algorithms

Cells with critically short telomere lengths are also known to undergo senescence once they approach their Hayflick limit [35], therefore we investigated the potential changes due to telomere length using the DNAm predictor for telomere length (DNAmTL) [36]. We found significant alterations in the DQ group, but not in the DQF group. Specifically, we observed a significant decrease in PC DNAm telomere length after the whole treatment (p-value=0.01, see Figure 3A), which was even more significant after adjusting by age (p-value=4.4·10^−4^, see Figure 3B). Importantly, the difference in telomere length acceleration between baseline and 3 months was larger and more significant (p-value=7.5·10^−5^) than the difference between baseline and 6 months (p-value=4.4·10^−4^).

**Figure 3.**
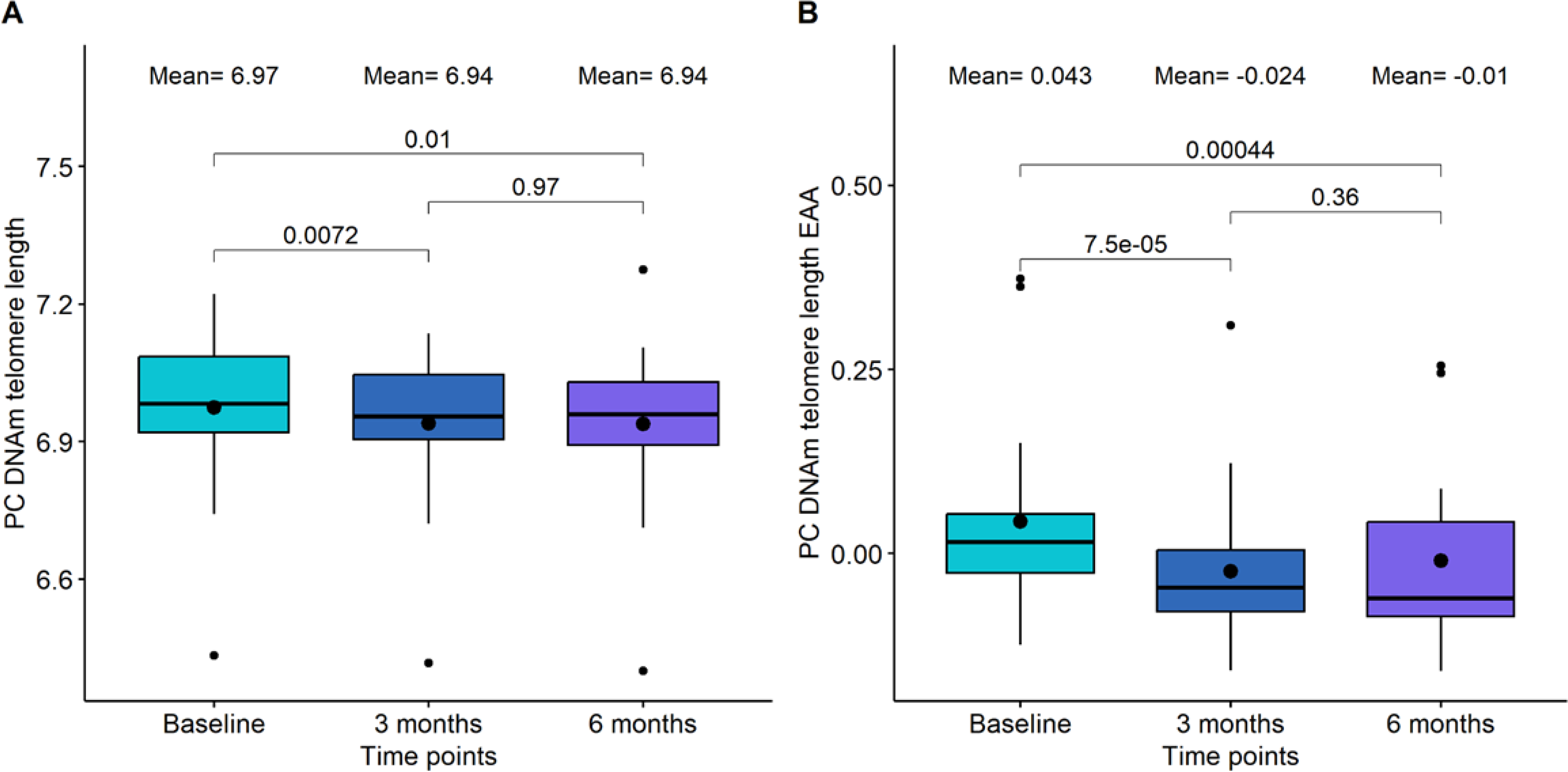
Boxplot showing the evolution of DNA methylation (DNAm) based telomere length in the DQ study. (A) DNAm telomere length. (B) DNAm telomere length acceleration. In the X-axis, the time points of measurements, in the Y axis, the epigenetic metric. On the top, the mean values at each time point. The p-values of the paired t-tests are also displayed in the plots.

Mitotic clock metrics were also employed to evaluate relative changes in stem cell replication. At the 3-month mark of DQ treatment, we observed a decrease in both the total number of stem cell divisions and the intrinsic tissue stem cell divisions, although these changes were not statistically significant (p-value=0.22 and p-value=0.22, respectively, see Figure 4). However, between 3-month and 6-month points, a significant increase was evident in both mitotic clocks (p-value=2.4·10^−4^ and p-value= 7.8·10^−4^, respectively). In the case of DQF treatment, no significant differences were found between the baseline and the 6-month measurement.

**Figure 4.**
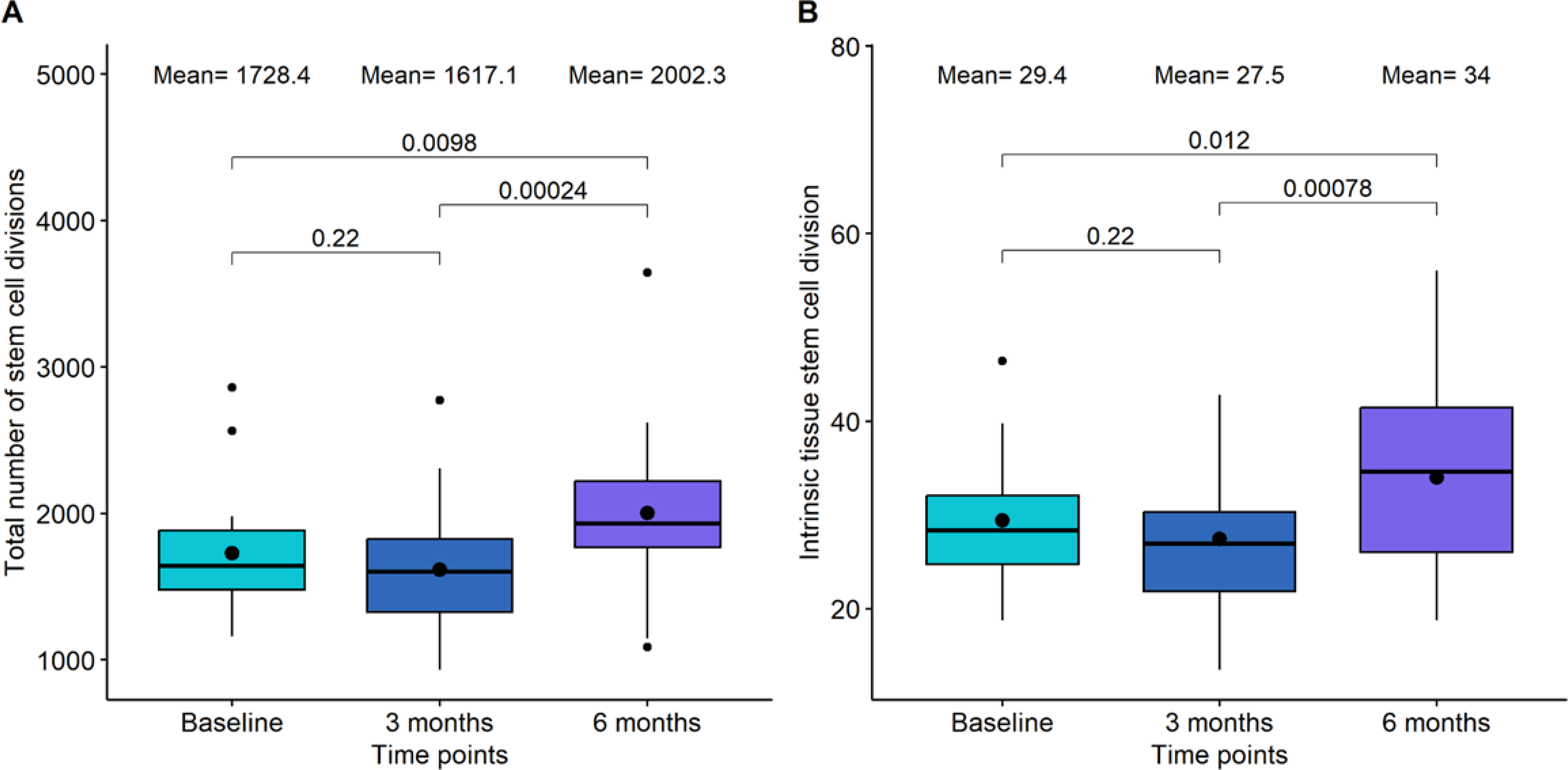
Boxplot showing the evolution of mitotic clocks in the Dasatinib and Quercetin study. (A) Total number of stem cell divisions. (B) Intrinsic tissue stem cell division. In the X-axis, the time points of measurements, in the Y axis, the number of divisions. On the top, the mean values at each time point. The p-values of the paired t-tests are also displayed in the plots.

### Impact of senolytic drugs on whole blood immune cell composition

We utilized EpiDISH (2023) to quantify 12 different immune cell subsets and assess changes within these subsets. During the 6-month period of DQ treatment, significant alterations were observed in CD4T Naive cells, B Naive cells, and monocytes (**Table 4**). The most notable and significant change was in CD4T Naive Cells, which exhibited a slight decrease at the 3-month mark (p-value=0.628) and experienced a more substantial decline between the 3 and the 6- month marks (p-value=0.029). B Naive cells displayed an insignificant increase for the global treatment (p-value=0.059), but we observed a significant increase between 3 and 6 months (p-value=0.001). Monocytes showed a global increase after 6 months of treatment (p-value=0.003) that was characterized by a significant decrease at 3 months (p-value=0.035) followed by a significant increase between 3 and 6 months (p-value=3.0·10^−5^). Conversely, CD4T Memory, CD8T Naive, CD8T Memory, B Memory, basophil, regulatory T cells, eosinophil, Natural Killer, and Neutrophil did not exhibit significant changes.

**Table 4.**
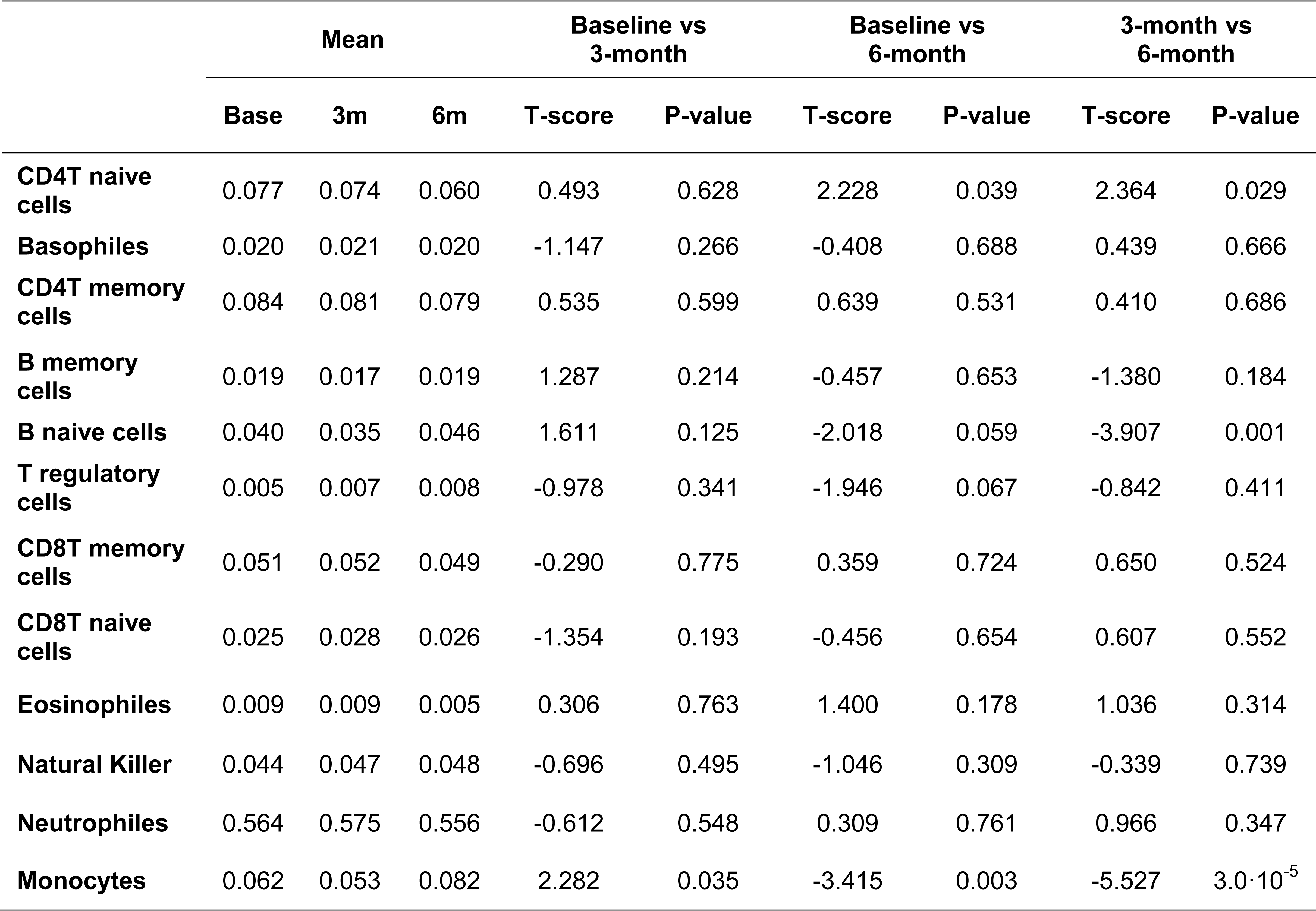
Statistical analysis for comparing baseline, 3 months, and 6 months immune cell proportions in the Dasatinib and Quercetin Study. The first three columns show the mean values for each immune cell proportion at each time point. The next columns have information about the t-test between baseline and 3-month test, between baseline and 6-month test, and between 3-month and 6- month tests, respectively.

Regarding the impact of DQF on immune cells, B Naive cells (Bnv) demonstrated a significant decrease after 6 months (p-value=3.0·10^−4^), which contrasts with the observations from DQ treatment. No significant changes were observed in the proportions of other immune cell subsets (**Table 5**).

**Table 5.**
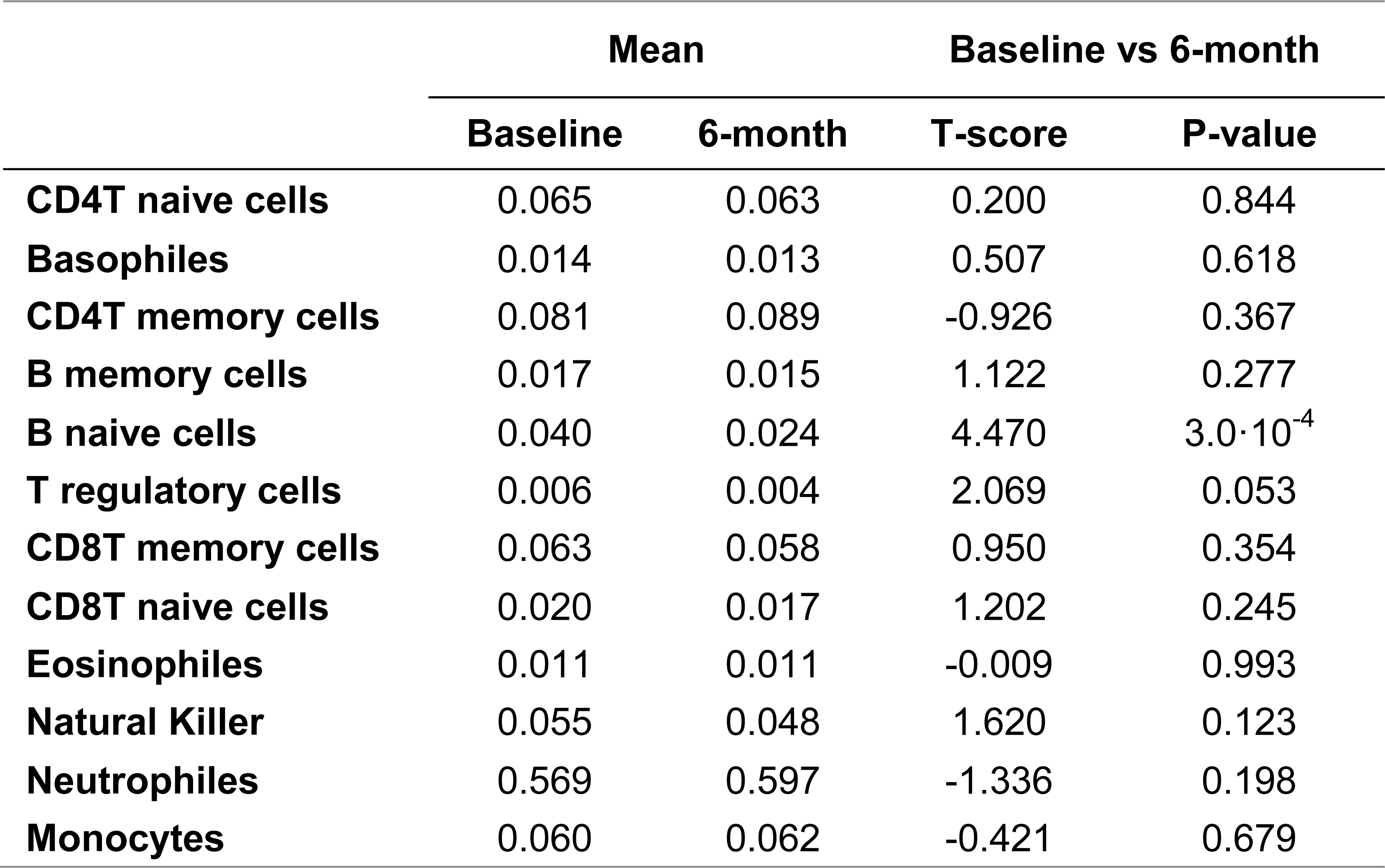
Statistical analysis for comparing baseline and 6-month immune cell proportions in the Dasatinib, Quercetin, and Fisetin Study. The first two columns show the mean values for each immune cell proportion at each time point. The next columns have information about the t-test between baseline and 6-month test.

Since most of the epigenetic clocks, especially the first-generation clocks, are dependent on immune subsets, we calculated the correlation between the EAA metrics and the immune cells proportions. As expected, we did not observe significant correlations between IntrinClock and immune cells. However, we observed high correlations between the other clocks and most of the immune cells (Figure 5). Thus, we decided to calculate immune EAA adjusting EAA values by all the immune cells that were significantly associated to the clocks (CD4T naive and memory cells, B naive and memory cells, CD8T naive and memory cells, natural killers, and neutrophils) and see whether the trends in first-generation clocks after DQ treatment were maintained. We found that the significance and direction of the associations were not modified after adjusting by immune cells, indicating that the increase of epigenetic age after DQ treatment was not due to the alteration of immune cell subsets (**Supplementary Figure S1**).

**Figure 5.**
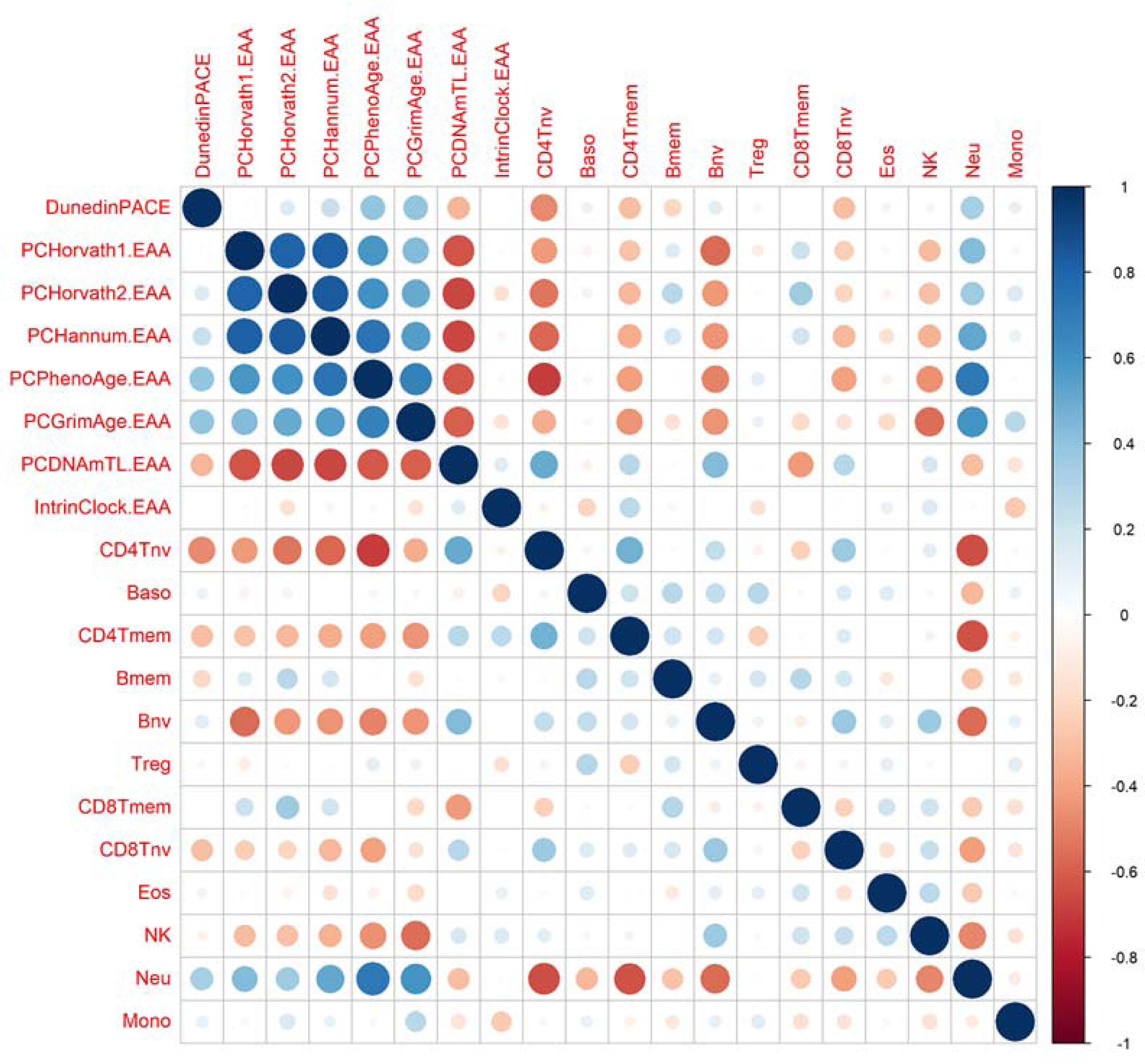
Correlation between epigenetic clocks and immune cell types. The size of the dots is proportional to the correlation value, being blue a positive correlation and red a negative correlation

### Impact of senolytic drugs on whole-genome DNA methylation

We also assessed global modification of DNAm in those individuals who were treated with DQ and those with DQF. To this end, we performed an Epigenome-wide association study (EWAS) comparing the methylation levels for all the CpG sites in the genome at different timepoints in each trial (**Supplementary Table S1**).

The first EWAS was performed between baseline and 3 months of DQ treatment. In this case, we identified 11 CpG sites differentially methylated, 4 of them hypermethylated and 7 hypomethylated after 3 months. These probes were mapped to 8 genes. Among them, *TGIF1, SORBS2,* and *ZNF768* were implicated in senescence [37]–[39]. Using a less restrictive threshold of p-value lower than 1·10^−4^, we performed an enrichment analysis. Among the 305 probes identified, we found three enriched processes highly related with senescence, such as glycolic process, vesicle recycling and endocytosis, and cytoskeletal organization [40]–[42].

Second, we evaluated the differences at global methylation between baseline and 6 months after DQ treatment. In this case, we only saw 2 CpG sites differentially methylated with an adjusted p-value lower than 0.05. One of them was hypermethylated and the other hypomethylated after a period of 6 months. The GREAT analysis was performed with the 475 CpG sites with a nominal p-value lower than 1·10^−4^. Although multiple gene ontology terms were identified as enriched, none of them were directly associated with aging or senescence.

Finally, when we compared the methylation levels between baseline and 6 months after DQF treatment, we identified 208 significant probes. Among them, approximately 50% were hypomethylated and 50% were hypermethylated. The GREAT analysis was performed using 556 probes and revealed multiple enriched pathways associated with senescence, such as epithelial cell proliferation, platelet dense granule membrane, cell junction, and positive regulation of cardiac muscle cell apoptotic process [43]–[47].

### Clinical and DNAm Proteomic Surrogate Analysis

The major hypothesized mechanism for the negative impacts of senescence is through the increased senescence-associated secretory phenotypes (SASP) which lead to high inflammatory cytokine signaling from senescent cells in a paracrine fashion. As an alternative to robust clinical lab measurements of inflammatory mediators, we used methylation risk scores surrogates to predict and quantify predicted changes in circulating proteomic markers [48]. The quantification of these markers and the comparison between the different timepoints are included in **Supplementary Table S2**).

We paid special attention to inflammation and inflammatory proteomic EpiScore analysis and these are listed in **Table 6**. In this analysis, we see some increased inflammatory mediators at 3 months which decrease from the 3 to 6-month timepoints. These inflammatory associated proteins include CRP, CXCL9, CXCL11, CCL17, and TGF-alpha. We see opposite trends with other inflammation associated markers such as Complement C4 and Complement C5a. Many inflammatory mediators for the innate adaptive immune system were not registered as significant in this analysis.

**Table 6.**
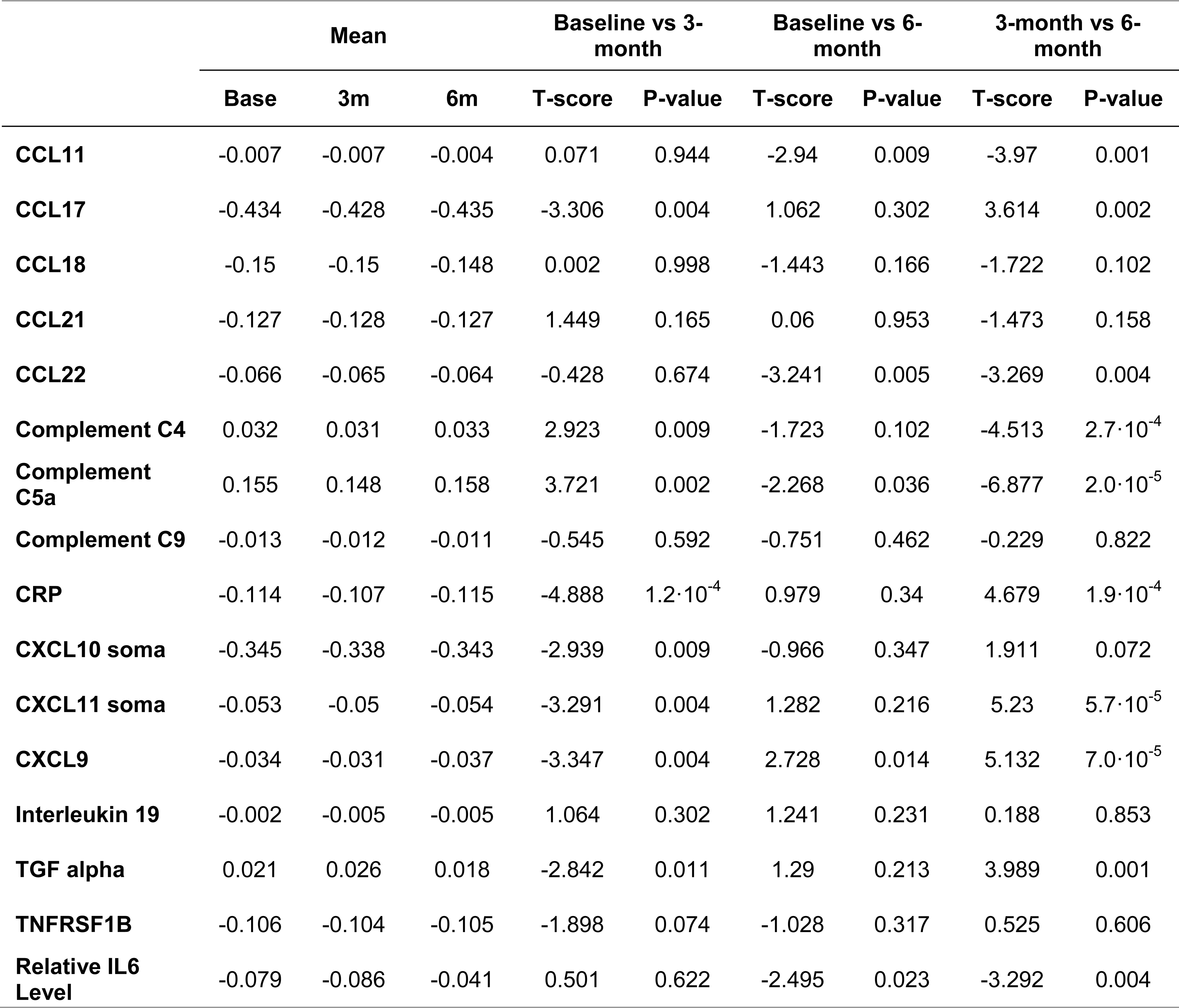
Inflammation and inflammatory proteomic EpiScore analysis between baseline, 3-month test, and 6-month test in the Dasatinib and Quercetin (DQ) trial. The first three columns show the mean values for each protein methylation risk score (MRS) at each time point. The next columns have information about the t-test between baseline and 3-month test, between baseline and 6-month test, and between 3-month and 6-month tests, respectively.

## Discussion

Overall, our findings indicate that the administration of senolytic drugs Dasatinib and Quercitin significantly increases biological age measured by first generation clocks. The only 2nd generation clock to show an increase was DNAmPhenoAge. However, we see no significant change in second and third-generation clocks such as GrimAge and DunedinPACE or any significant changes with the addition of Fisetin to the protocol (DQF treatment).

The increase in DNAmPhenoAge is different in trend from the other phenotypically trained clocks but is not surprising as this clock has previously been described as a hybrid between first and second-generation clocks. Unlike GrimAge and DunedinPoAm, the dependent variable used for training DNAmPhenoAge, also included chronological age. While GrimAge included observed age in its derivation on the other side of the equation—as a predictor along with DNAm—essentially adjusting out its effects. This similarity is further described by its module composition which reflects similarity to first generation trained chronological clocks [49].

The biological aging impacts effects of this analysis are difficult to analyze. While, the link between cellular senescence and aging is indisputable [50]. The differential analysis of these clocks limits our interpretation of their clinical significance.

Previous studies have shown that the first-generation clocks, such as the Skin&blood clock, will increase with cell passage and time in cell culture. This increase was shown irrespective of cells which have human telomerase reverse transcriptase (hTERT) expressed and thus didn’t enter replicative senescence or have telomere attrition which suggest that these first-generation clocks are measuring a process separate from either senescence or telomere attrition [51]. This suggests that epigenetic age clocks might not do a good job in measuring the process of senescence or the biological impact of senescence.

However, the non-significant change in 2nd and 3rd generation clocks such as GrimAge and DunedinPACE is also informative. 2nd and 3rd generation clocks, trained to phenotypes of aging, are more prone to capture the underlying biology of aging since these are trained to biomarkers of the biological aging process and not just correlating CpGs to chronological age. It is plausible that the significant biological age increases that are seen in the first-generation clocks, and DNAmPhenoAge, might be accelerated due to the age correlated CpG locations and not the underlying biological relevant impacts since the more predictive, and more biological associated, 2^nd^ and 3^rd^ generation clocks are not showing the same increase. Thus, while neither generation of clocks are showing improvement with senolytic treatment, the increases in the 1^st^ generation clocks are less reliable for predicting the phenotypic outcomes and thus should be interpreted carefully.

This ability for different generations of clocks to reflect different interventional change is not new. In previous DNAm interventional studies such as the CALERIE study, we see that caloric restriction only displayed significant aging changes with DunedinPACE, yet didn’t show significance with any other clock [52].

Another confounding variable which limits our ability to establish biological aging significance is the significant changes in CD4T and CD8T Naive cells, B Naive cells, and monocytes. Immune cell changes with aging have been a large confounding error in previous DNAm clocks. For instance, previous studies have shown that human naive CD8+ T cells can exhibit an epigenetic age 15–20 years younger than effector memory CD8+ T cells from the same individual. This means that previous epigenetic clocks measure two independent variables, aging and immune cell composition [53]. To analyze if immune changes were responsible for the first-generation epigenetic clock acceleration, we calculated immune EAA and adjusted by all the immune cells that were significantly associated to the clocks (CD4T naive and memory cells, B naive and memory cells, CD8T naive and memory cells, natural killers, and neutrophils). We found that the significance and direction of the associations were not modified after adjusting by immune cells, indicating that the increase of epigenetic age after DQ treatment was not due to the modulation of immune cell subsets but representing an increase due to the CpG inclusion and weights of the clocks themselves.

Furthermore, we also analyzed the IntrinClock to assess the relationship of immune cell subtypes. The IntrinClock was created to be independent of immune cell subset changes. And created through an analysis of the age-related changes in independent cell types. This clock showed no significant change in any of the treatment arms. This might be due to the unique construction method for the IntrinClock. IntrinClock was generated via the deliberate removal of CpGs that were characteristic of naïve cells. It is possible that senolytic treatment is affecting a subset of CpGs that were removed in the construction of the IntrinClock that are more correlated to general properties of naïve cells (quiescence, etc.) rather than those that represent immune cell type composition specifically.

The major hypothesized mechanism for the negative impacts of senescence is through the increased senescence-associated secretory phenotypes (SASP) which lead to high inflammatory cytokine signaling from senescent cells in a paracrine fashion. Although SASP proteins were not measured directly in this study, we used DNA methylation risk scores for protein surrogates to analyze changes in common SASP proteins. In some cases, these methylation risk scores have been shown to have better resolution and connection to outcomes than traditional measures. For instance, the Episcore for C-Reactive protein has shown age-related associations in cohorts which were not seen with log(CRP) clinical measures and association to cognitive function and brain MRIs [54]. Looking at these methylation risk scores in our longitudinal data, we see some interesting trends of increased inflammatory mediators at 3 months which decrease from the 3-6 month timepoints. These inflammatory associated proteins include CRP, CXCL9, CXCL11, CCL17, and TGF-alpha. We see opposite trends with other inflammation associated markers such as Complement C4 and Complement C5a. The accuracy of the EpiScore protein predictions to measured proteins is still low. Thus, the changes we see here are limited by accuracy of the prediction. CRP, which is the most validated of the proteomic EpiSign scores, shows decreases at 3 months and increases at 6 months which might suggest duration of senolytic therapy might impact the phenotypic outcomes. Furthermore, the treatment impacts on the SASP might also indicate a plausible explanation for the differences in age accelerations between the first generation IntrinClock and other first-generation clocks. SASP-related CpG sites would be differentially methylated in naïve T cells and thus removed from construction of the IntrinClock but not Horvath, Hannum, or PhenoAge. Clocks.

Together, our findings suggest that 1st generation clocks are insufficient tools to quantify the impact of senolytic therapies. Additionally, second generation clocks still remain largely unchanged with Dasatinib and Quercetin use and other markers are needed to measure the physiologic impact of senolytic treatments. In either instance, our findings reemphasize the importance of developing new biomarkers which can quantify senescence and impact to aging phenotypes.

Strengths of the present study include its prospective longitudinal design, the duration of longitudinal measures, the standardized and batch normalized epigenetic data, and comprehensive inclusion of novel and complementary epigenetic age measures that were repeatedly collected.

There are also some important limitations. First, was the limitation of small sample size and the lack of a control group. Having epigenetic methylation changes in the control group and a large sample size may have improved statistical power to detect small effects. Likewise, the sample size did not allow a rigorous analysis of individual CpG sites with correction for multiple comparisons. Future, larger studies should replicate the present results and extend them by examining other epigenetic clocks and individual CpG sites with appropriate corrections for multiple testing.

Secondly, epigenetic aging markers were not supplemented with classical blood measurements or classical phenotypic markers of aging and thus, it is possible that some unmeasured confounders biased our results. However, age acceleration was independent of potential confounders including chronological age and sex.

Finally, DNA methylation was measured in blood samples only. As senescence is different in every tissue type, blood based DNAm data might not capture the physiologic impact of senolytic therapy and future studies in tissue specific analysis might provide more insight.

Future studies can improve on our limitations by including large sample size, additional clinical biomarkers of inflammation and senescence, and incorporate analysis into tissues more associated with senescence related aging phenotypes. We hope this data can be reanalyzed as new senolytic DNA methylation analysis tools become available.

## Materials and methods

### Study participants and senolytic administration

For the evaluation of DQ treatment upon epigenetic age, 19 study participants were accrued from November 2020 to December 2020 at the Institute for Hormonal Balance, Orlando. **Table 1** shows the demographic and clinical characteristics of these participants. Adults aged 40 and older able to comply with treatment plan and laboratory tests were included. Individuals with neoplastic cancer within 5 years prior to screening, immune disease, viral illness, cardiovascular or cerebrovascular disease, ischemic attack in the last 6 months, hepatitis or HIV, Body mass index higher than 40kg/m2, active infection, or previously used DQ were excluded. Informed consent was obtained from study participants. The FDA registered IRB (Institute for Regenerative and Cellular Medicine) approved this study, which is registered at ClinicalTrials.gov (NCT04946383) and which is an ongoing clinical trial to determine the effectiveness of Quercetin and Dasatinib supplements on the patient’s epigenetic aging rate. The treatment comprised 500mg Quercetin and 50mg Dasatinib oral capsules on Monday, Tuesday, and Wednesday (3 days in a row) per month for the duration of 6 months. It is worth mentioning that three subjects stopped the treatment after 3 months due to nausea, prostate cancer diagnosis, and concern about the drug, respectively.

For the evaluation of DQF treatment, all the participants from the first study were invited to join and new participants were recruited from June 2022 to July 2022 at the Institute for Hormonal Balance, Orlando. From the previous study, 10 participants joined this study, and 9 participants were newly recruited. The same inclusion and exclusion criteria were followed as in the DQ study. In this case, the treatment consisted of the same dosage and timeline as in the DQ treatment but included 500 mg of Fisetin oral capsules on Monday, Tuesday, and Wednesday (3 days in a row) per month for the duration of 6 months. Moreover, 8 participants got a strawberry based Fisetin and 11 got a non-strawberry based Fisetin.

### DNA methylation assessment

Peripheral whole blood samples were obtained using the lancet and capillary method and immediately mixed with lysis buffer to preserve the cells. DNA extraction was performed, and 500 ng of DNA was subjected to bisulfite conversion using the EZ DNA Methylation kit from Zymo Research, following the manufacturer’s protocol. The bisulfite-converted DNA samples were then randomly allocated to designated wells on the Infinium HumanMethylationEPIC BeadChip. The samples were amplified, hybridized onto the array, and subsequently stained. After washing steps, the array was imaged using the Illumina iScan SQ instrument to capture raw image intensities, enabling further analysis.

*Minfi* R package was used for the pre-processing of DNAm data. We pre-processed all the samples from the different studies together to remove batch effects. In the sample quality control, we removed those samples with aberrant methylation levels and with background signal levels (mean p-value higher than 0.05). We also discarded those probes with background signal following the same threshold. We further normalized the methylation values using the Genome-wide Median Normalization (GMQN) and the Beta Mixture Quantile (BMIQ) methods. Finally, we imputed the missing values using the k-nearest neighbors (knn) algorithm. Finally, we used a 12-cell immune deconvolution method developed by Zhend et al to estimate cell type proportions. We chose this method because in previous analysis we saw this has R^2^ of 0.96 and above to immune cell subsets measured by RNA-seq and flow cytometry [55].

### Statistical analyses and reproducibility

#### DNA methylation clocks and related measures

We used DNAm data to calculate a series of measures broadly known as epigenetic clocks. We computed four clocks designed to predict the chronological age of the donor, Horvath Pan Tissue, Horvath Skin and Blood, and Hannum; two clocks designed to predict mortality, the DNAmPhenoAge and GrimAge clocks; a clock to measure telomere length, DNAmTL [36]; two clock designed to measure mitotic age, total stem cell divisions (tnsc) and intrinsic tissue stem cell division (irS) [56]; a DNAm measure of the rate of deterioration in physiological integrity, the DundedinPACE; and a clock to measure chronological age but not dependent on immune cells, the IntrinClock.

To calculate the principal component-based epigenetic clock for the Horvath multi-tissue clock, Hannum clock, DNAmPhenoAge clock, GrimAge clock, and telomere length we used the custom R script available via GitHub (https://github.com/MorganLevineLab/PC-Clocks). Non- principal component-based (non-PC) Horvath, Hannum, and DNAmPhenoAge epigenetic metrics were calculated using the *methyAge* function in the *ENMix* R package. The pace of aging clock, DunedinPACE, was calculated using the *PACEProjector* function from the *DunedinPACE* package available via GitHub (https://github.com/danbelsky/DunedinPACE). The mitotic clocks were calculated using the *epiTOC2* function from the *meffonym* package. Finally, the IntrinClock was calculated as described elsewhere [53].

To calculate the EAA of multiple clocks, we fit a regression model between the chronological age of the individuals and the different epigenetic age measures. We also included the batch PCs as a fixed effect as a way to control for potential batch effects. This methodology of incorporating PCs in EAA calculations was previously described in Joyce et al.

#### Differentially methylated loci analysis

The epigenome-wide association study (EWAS) was performed using the *limma* Bioconductor package. We performed a differential mean analysis on different timepoints to see whether the treatment was associated with changes at specific loci. Based on the available covariates, we adjusted all the regression models by sex, age, batch effect, and three principal components. We also set as random effect the participant ID. For each timepoint comparison, we fitted models

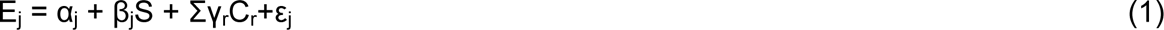

where E_j_ denotes the methylation level vector across individuals at probe j (j = 1, . . . 866836), S is the time point with its associated effect, β_j_, C_r_ is the r adjusting covariate and its effect γ_r_, and ε_j_ is the noise that follows the distribution of methylation levels with mean 0. Adjusted P-values were calculated using FDR correction for considering multiple comparisons. The inflation or deflation of P-values across the methylome was assessed with Q-Q plots and lambda values. We selected as significant probes those with FDR lower than 0.05 after correcting for multiple comparisons.

We next used GREAT to understand the functional relevance of the differentially methylated loci (DML) with a nominal p-value lower than 1·10^−4^. The GREAT software will compare genomic features against the genes of interest in order to run Gene Ontology (GO) analysis. This software looks at the number of DMLs which overlap to the promoter and enhancer regions to run a binomial enrichment analysis of identifying overrepresented/enriched GO terms.

## Author contributions

EL performed patient recruitment, clinical management, and sample procurement. NC and VD performed methylation preprocessing, data analysis, and statistical analysis. EV and AT analyzed IntrinClock data. LN and MC provided immune cell subset analysis. TM conducted methylation laboratory analysis. HW, AL, LT, LL, and RS helped with study design, manuscript drafting, and submission.

## Data Availability

All data produced in the present study are available upon reasonable request to the authors.

## Acknowledgements

We are grateful to all participants and researchers who took part in this study.

## Conflicts of interest

NC, VBD, RS, HW, AL, LT, and TLM are employees of TruDiagnostic.

## Ethical Statement

The study involving human participants was reviewed and approved by the Institute for Regenerative and Cellular Medicine. The participants provided informed consent to participate in this study.

## Funding

TruDiagnostic has provided funding for data analysis and IRB funding. The Institute for Hormonal Balance provided all other costs associated with testing and patient recruitment.

## Notes

### Clinical Trial

NCT04946383

### Author Declarations

The FDA registered IRB (Institute for Regenerative and Cellular Medicine) approved this study. The participants provided informed consent to participate in this study.

