## Supplementary Materials for "Exploring the effects of Dasatinib, Quercetin, and Fisetin on DNA methylation clocks: a longitudinal study on senolytic interventions"

### Figures

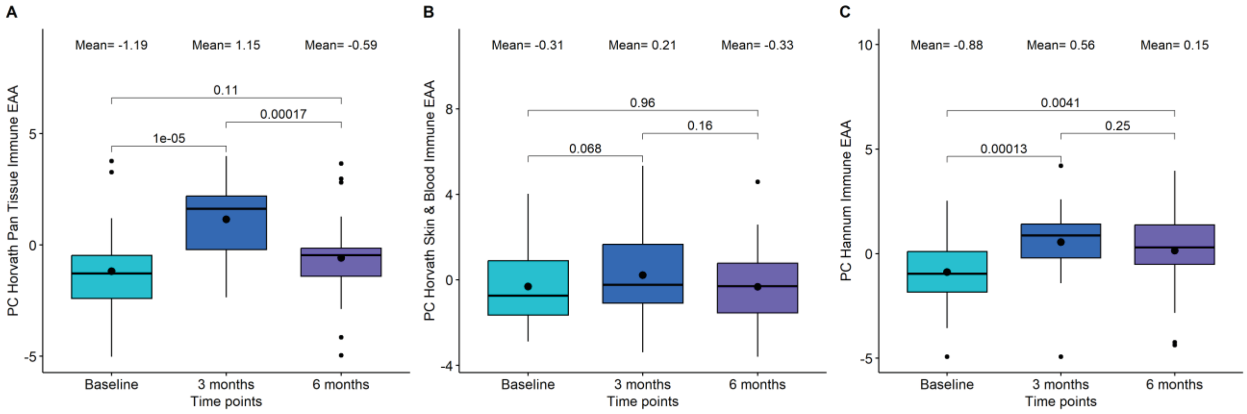


**Figure S1. Boxplot showing the evolution of epigenetic age acceleration (EAA) first-generation clocks after adjusting by immune cells in the Dasatinib and Quercetin (DQ) study.** (A) PC Horvath pan tissue Immune EAA. (B) PC Horvath Skin&Blood Immune EAA. (C) PC Hannum Immune EAA. In the X-axis, the time points of measurements, in the Y axis, the EAA measure adjusted by immune cells. On the top, the mean values at each time points. The p-values of the paired t-tests are also displayed in the plots.

### Tables

**Table S1. Differentially methylated loci for Dasatinib and Quercetin (DQ) treatment and Dasatinib, Quercetin, and Fisetin (DQF) treatment.** All the significant probes (FDR<0.05) for DQ analyses are included. However, only the top 15 probes are included from the DQF analysis.

| **Dasatinib and Quercetin** | | | | | |
|---|---|---|---|---|---|
| **0-3 months** | | | | | |
| **CpG site** | **logFC** | **diffMeth** | **P.Value** | **FDR** | **geneID** |
| cg05915794 | 0.2878 | Hyper | $1.5 \cdot 10^{-12}$ | $1.3 \cdot 10^{-6}$ | HRNBP3 |
| cg18889307 | -0.08974 | Hypo | $6.9 \cdot 10^{-9}$ | 0.002 | TGIF1 |
| cg02889828 | -0.1833 | Hypo | $8.6 \cdot 10^{-9}$ | 0.002 | |
| cg27262015 | -0.1243 | Hypo | $2.0 \cdot 10^{-8}$ | 0.004 | SORBS2 |
| cg04704414 | 0.1107 | Hyper | $1.3 \cdot 10^{-7}$ | 0.022 | NUBP2 |
| cg06902281 | 0.1175 | Hyper | $1.9 \cdot 10^{-7}$ | 0.027 | ZNF169 |
| cg04955246 | -0.155 | Hypo | $2.5 \cdot 10^{-7}$ | 0.031 | PRKCA |
| cg02840515 | -0.1262 | Hypo | $4.3 \cdot 10^{-7}$ | 0.039 | KIAA2012 |
| cg04132292 | -0.2873 | Hypo | $4.4 \cdot 10^{-7}$ | 0.039 | |
| cg25024143 | -0.1463 | Hypo | $4.6 \cdot 10^{-7}$ | 0.039 | SP2 |
| cg20273352 | -0.1645 | Hypo | $5.6 \cdot 10^{-7}$ | 0.044 | IGHMBP2 |
| **0-6 months** | | | | | |
| **CpG site** | **logFC** | **diffMeth** | **P.Value** | **FDR** | **geneID** |
| cg10585661 | -0.1102 | Hypo | $8.0 \cdot 10^{-8}$ | 0.035 | FAM131A |
| cg05779406 | 0.271 | Hyper | $8.1 \cdot 10^{-8}$ | 0.035 | ZFAND2A |
| **Dasatinib, Quercetin, and Fisetin** | | | | | |
| **0-6 months** | | | | | |
| **CpG site** | **logFC** | **diffMeth** | **P.Value** | **FDR** | **geneID** |
| cg00379708 | -0.1492 | Hypo | $1.4 \cdot 10^{-9}$ | $6.7 \cdot 10^{-4}$ | RBM20 |
| cg13172627 | 0.5832 | Hyper | $1.9 \cdot 10^{-9}$ | $6.7 \cdot 10^{-4}$ | VENTXP1 |
| cg26676360 | 0.1803 | Hyper | $2.3 \cdot 10^{-9}$ | $6.7 \cdot 10^{-4}$ | LRP1 |
| cg02632099 | 0.3497 | Hyper | $3.5 \cdot 10^{-9}$ | $7.5 \cdot 10^{-4}$ | |
| cg18054725 | -0.1509 | Hypo | $9.2 \cdot 10^{-9}$ | 0.002 | ICAM3 |
| cg01613010 | 0.2131 | Hyper | $2.5 \cdot 10^{-8}$ | 0.002 | |

| | | | | | |
|---|---|---|---|---|---|
| cg16640929 | -0.6613 | Hypo | $2.6 \cdot 10^{-8}$ | 0.002 | ZBTB12 |
| cg00877151 | 0.4233 | Hyper | $2.8 \cdot 10^{-8}$ | 0.002 | IPO11-LRRC70;IPO11 |
| cg06310713 | 0.3591 | Hyper | $3.1 \cdot 10^{-8}$ | 0.002 | PALLD |
| cg08187458 | 0.1809 | Hyper | $3.3 \cdot 10^{-8}$ | 0.002 | |
| cg00288652 | -0.1792 | Hypo | $3.5 \cdot 10^{-8}$ | 0.002 | |
| cg04706867 | -0.1987 | Hypo | $3.6 \cdot 10^{-8}$ | 0.002 | CNTNAP2 |
| cg09093137 | -0.5388 | Hypo | $3.7 \cdot 10^{-8}$ | 0.002 | SRPK2 |
| cg03012879 | -0.4304 | Hypo | $3.7 \cdot 10^{-8}$ | 0.002 | HMGCR |
| cg12993163 | -0.1746 | Hypo | $4.2 \cdot 10^{-8}$ | 0.002 | SHOX2 |

**Table S2. Statistical analysis for comparing baseline, 3 month, and 6 month Marioni markers proportions in the Dasatinib and Quercetin Study.** The first three columns show the mean values for each immune cell proportion at each time point. The next columns have information about the t-test between baseline and 3-month test, between baseline and 6-month test, and between 3-month and 6-month tests, respectively.

| | Mean | | | Baseline vs 3-month | | Baseline vs 6-month | | 3-month vs 6-month | |
|---|---|---|---|---|---|---|---|---|---|
| | **Base** | **3m** | **6m** | **T-score** | **P-value** | **T-score** | **P-value** | **T-score** | **P-value** |
| **Epigenetic. Age..Zhang.** | -1.863 | -1.863 | -1.405 | -0.001 | 0.999 | -4.964 | 0.0001 | -4.373 | $3.7 \cdot 10^{-4}$ |
| **Alcohol** | -11.698 | -11.752 | -11.61 | 0.746 | 0.465 | -0.853 | 0.405 | -1.616 | 0.123 |
| **Body Fat** | -10.065 | -10.662 | -11.028 | 1.177 | 0.255 | 2.221 | 0.039 | 0.882 | 0.389 |
| **Body Mass Index** | -0.662 | -0.687 | -0.65 | 1.8 | 0.089 | -0.78 | 0.446 | -2.53 | 0.021 |
| **HDL Cholesterol** | 2.644 | 2.682 | 2.669 | -1.158 | 0.262 | -0.894 | 0.383 | 0.482 | 0.636 |
| **Smoking** | 2.809 | 2.786 | 2.764 | 0.688 | 0.5 | 1.087 | 0.291 | 0.568 | 0.577 |
| **Waist Hip Ratio** | -0.336 | -0.338 | -0.332 | 0.767 | 0.453 | -1.116 | 0.279 | -1.84 | 0.082 |
| **ADAMTS** | 0.111 | 0.109 | 0.11 | 1.36 | 0.191 | 0.92 | 0.37 | -0.563 | 0.58 |
| **Adiponectin** | -0.063 | -0.057 | -0.066 | -4.033 | 0.001 | 3.237 | 0.005 | 5.362 | $4.3 \cdot 10^{-5}$ |
| **Afamin** | -0.009 | -0.009 | -0.009 | -0.797 | 0.436 | -0.869 | 0.396 | -0.213 | 0.834 |
| **Alpha Liduronidase** | 0.112 | 0.115 | 0.111 | -5.149 | $6.7 \cdot 10^{-5}$ | 1.031 | 0.316 | 5.468 | $3.4 \cdot 10^{-5}$ |
| **Aminoacylase.1** | -0.336 | -0.339 | -0.333 | 1.975 | 0.064 | -1.261 | 0.224 | -2.651 | 0.016 |
| **B2.microglobulin** | -0.373 | -0.367 | -0.373 | -2.831 | 0.011 | -0.145 | 0.886 | 2.633 | 0.017 |

| | | | | | | | | | |
|---|---|---|---|---|---|---|---|---|---|
| **BMP.1** | 0.121 | 0.121 | 0.118 | -0.039 | 0.969 | 2.045 | 0.056 | 2.55 | 0.02 |
| **CCL11** | -0.007 | -0.007 | -0.004 | 0.071 | 0.944 | -2.94 | 0.009 | -3.97 | 0.001 |
| **CCL17** | -0.434 | -0.428 | -0.435 | -3.306 | 0.004 | 1.062 | 0.302 | 3.614 | 0.002 |
| **CCL18** | -0.15 | -0.15 | -0.148 | 0.002 | 0.998 | -1.443 | 0.166 | -1.722 | 0.102 |
| **CCL21** | -0.127 | -0.128 | -0.127 | 1.449 | 0.165 | 0.06 | 0.953 | -1.473 | 0.158 |
| **CCL22** | -0.066 | -0.065 | -0.064 | -0.428 | 0.674 | -3.241 | 0.005 | -3.269 | 0.004 |
| **CCL25.C.C** | -0.064 | -0.062 | -0.063 | -1.632 | 0.12 | -0.698 | 0.494 | 1.138 | 0.27 |
| **CD163** | -0.242 | -0.239 | -0.243 | -1.699 | 0.107 | 0.449 | 0.659 | 2.327 | 0.032 |
| **CD209.antigen** | 0.13 | 0.126 | 0.129 | 2.813 | 0.012 | 1.143 | 0.268 | -1.696 | 0.107 |
| **CD48.antigen** | -0.152 | -0.148 | -0.154 | -2.497 | 0.022 | 1.457 | 0.162 | 4.427 | 0.00033 |
| **CD6** | 0.098 | 0.094 | 0.101 | 1.717 | 0.103 | -0.798 | 0.435 | -2.638 | 0.017 |
| **CDL5** | -0.069 | -0.063 | -0.067 | -2.946 | 0.009 | -1.482 | 0.156 | 2.037 | 0.057 |
| **CHIT.1** | -0.169 | -0.165 | -0.168 | -3.103 | 0.006 | -0.608 | 0.551 | 1.806 | 0.088 |
| **CLEC11A.e1** | -0.022 | -0.019 | -0.02 | -0.931 | 0.364 | -0.755 | 0.46 | 0.057 | 0.955 |
| **CLEC11A.e2** | -0.16 | -0.156 | -0.157 | -1.64 | 0.118 | -0.988 | 0.336 | 0.397 | 0.696 |
| **Coagulation.factor.VII** | 0.043 | 0.041 | 0.041 | 3.135 | 0.006 | 1.537 | 0.142 | -0.111 | 0.913 |
| **Complement.C4** | 0.032 | 0.031 | 0.033 | 2.923 | 0.009 | -1.723 | 0.102 | -4.513 | 0.00027 |
| **Complement.C5a** | 0.155 | 0.148 | 0.158 | 3.721 | 0.002 | -2.268 | 0.036 | -6.877 | 0.000002 |
| **Complement.c9** | -0.013 | -0.012 | -0.011 | -0.545 | 0.592 | -0.751 | 0.462 | -0.229 | 0.822 |
| **Contactin.4** | 0.171 | 0.169 | 0.17 | 1.494 | 0.153 | 0.62 | 0.543 | -0.608 | 0.551 |
| **CRP** | -0.114 | -0.107 | -0.115 | -4.888 | 0.00012 | 0.979 | 0.34 | 4.679 | 0.00019 |
| **CRTAM** | 0.057 | 0.056 | 0.057 | 0.583 | 0.567 | -0.396 | 0.696 | -0.928 | 0.366 |
| **CXCL10** | 0.138 | 0.137 | 0.137 | 0.289 | 0.776 | 0.272 | 0.789 | 0.09 | 0.93 |
| **CXCL10.soma** | -0.345 | -0.338 | -0.343 | -2.939 | 0.009 | -0.966 | 0.347 | 1.911 | 0.072 |
| **CXCL11** | 0.083 | 0.082 | 0.084 | 0.959 | 0.35 | -0.908 | 0.376 | -1.991 | 0.062 |
| **CXCL11.soma** | -0.053 | -0.05 | -0.054 | -3.291 | 0.004 | 1.282 | 0.216 | 5.23 | 0.000057 |
| **CXCL9** | -0.034 | -0.031 | -0.037 | -3.347 | 0.004 | 2.728 | 0.014 | 5.132 | 0.00007 |
| **E.selectin** | -0.031 | -0.033 | -0.031 | 1.922 | 0.071 | -0.051 | 0.96 | -2.244 | 0.038 |
| **Ectodysplasin.A** | -0.215 | -0.215 | -0.214 | -0.637 | 0.532 | -1.007 | 0.327 | -0.479 | 0.638 |
| **EN.RAGE** | 0.05 | 0.054 | 0.047 | -2.309 | 0.033 | 0.955 | 0.352 | 3.166 | 0.005 |
| **ENPP7** | -0.028 | -0.032 | -0.025 | 1.354 | 0.192 | -0.683 | 0.503 | -1.889 | 0.075 |
| **ESM.1** | -0.241 | -0.241 | -0.241 | -0.38 | 0.708 | -0.784 | 0.443 | -0.169 | 0.868 |
| **EZR** | -0.019 | -0.022 | -0.019 | 2.887 | 0.01 | 0.248 | 0.807 | -2.392 | 0.028 |
| **FAP** | -0.141 | -0.144 | -0.139 | 2.153 | 0.045 | -1.27 | 0.22 | -3.976 | 0.001 |
| **FCER2** | -0.271 | -0.273 | -0.261 | 1.193 | 0.248 | -6.205 | 0.0000074 | -5.795 | 0.000017 |
| **FCGR3A** | -0.199 | -0.192 | -0.202 | -4.338 | 0.0004 | 1.584 | 0.131 | 6.021 | 0.000011 |

| | | | | | | | | | |
|---|---|---|---|---|---|---|---|---|---|
| **FcRL2** | -0.176 | -0.176 | -0.174 | -0.737 | 0.47 | -2.343 | 0.031 | -1.875 | 0.077 |
| **FGF.21** | -0.117 | -0.115 | -0.119 | -0.811 | 0.428 | 1.55 | 0.139 | 2.454 | 0.025 |
| **G.CSF** | -0.01 | -0.01 | -0.012 | -0.416 | 0.682 | 0.553 | 0.587 | 0.998 | 0.332 |
| **Galectin.4** | -0.211 | -0.211 | -0.211 | 0.221 | 0.828 | -0.48 | 0.637 | -0.936 | 0.362 |
| **GDF.8** | 0.139 | 0.135 | 0.142 | 4.81 | 0.00014 | -2.63 | 0.017 | -6.549 | 0.00000 37 |
| **GHR** | 0.138 | 0.134 | 0.136 | 2.021 | 0.058 | 1.276 | 0.218 | -0.897 | 0.382 |
| **GPIba** | -0.358 | -0.355 | -0.353 | -1.564 | 0.135 | -3.273 | 0.004 | -1.197 | 0.247 |
| **Granulysin** | -0.12 | -0.12 | -0.121 | -0.143 | 0.888 | 0.359 | 0.724 | 0.494 | 0.627 |
| **Granzyme.A** | -0.031 | -0.032 | -0.031 | 0.394 | 0.698 | -0.147 | 0.885 | -0.553 | 0.587 |
| **GZMA** | -0.068 | -0.068 | -0.067 | -0.165 | 0.87 | -0.441 | 0.664 | -0.268 | 0.792 |
| **HCII** | 0.044 | 0.042 | 0.045 | 1.24 | 0.231 | -1.407 | 0.176 | -1.803 | 0.088 |
| **HGF** | 0.044 | 0.046 | 0.043 | -1.772 | 0.093 | 0.845 | 0.409 | 2.847 | 0.011 |
| **HGFA** | 0.359 | 0.359 | 0.358 | -0.212 | 0.835 | 0.38 | 0.708 | 0.642 | 0.529 |
| **HGFI** | 0.551 | 0.545 | 0.547 | 0.809 | 0.429 | 0.614 | 0.547 | -0.46 | 0.651 |
| **ICAM5** | -0.09 | -0.095 | -0.091 | 3.234 | 0.005 | 0.291 | 0.774 | -3.012 | 0.007 |
| **IGFBP.1** | -0.107 | -0.102 | -0.109 | -4.805 | 0.00014 | 2.733 | 0.014 | 8.189 | 0.00000 018 |
| **IGFBP.4** | -0.051 | -0.049 | -0.049 | -1.576 | 0.132 | -1.318 | 0.204 | 0.278 | 0.784 |
| **Insulin.rece ptor** | -0.136 | -0.136 | -0.135 | -0.39 | 0.701 | -1.276 | 0.218 | -0.98 | 0.34 |
| **Interleukin. 19** | -0.002 | -0.005 | -0.005 | 1.064 | 0.302 | 1.241 | 0.231 | 0.188 | 0.853 |
| **L.selectin** | 0.066 | 0.065 | 0.064 | 0.554 | 0.587 | 1.201 | 0.245 | 0.926 | 0.366 |
| **LFT** | -0.002 | -0.002 | -0.002 | 0.174 | 0.864 | -0.386 | 0.704 | -0.506 | 0.619 |
| **LGALS3BP** | 0.012 | 0.008 | 0.011 | 3.318 | 0.004 | 0.429 | 0.673 | -3.431 | 0.003 |
| **LY9** | -0.12 | -0.119 | -0.123 | -0.571 | 0.575 | 1.597 | 0.128 | 2.718 | 0.014 |
| **Lymphotoxi n.abeta** | -0.04 | -0.046 | -0.04 | 3.568 | 0.002 | -0.267 | 0.793 | -3.858 | 0.001 |
| **MIA** | 0.103 | 0.102 | 0.105 | 1.276 | 0.218 | -2.177 | 0.043 | -3.517 | 0.002 |
| **MMP.1.1** | -0.116 | -0.112 | -0.119 | -1.884 | 0.076 | 1.631 | 0.12 | 3.338 | 0.004 |
| **MMP.12** | -0.199 | -0.196 | -0.197 | -1.898 | 0.074 | -1.588 | 0.13 | 0.391 | 0.7 |
| **MMP.9** | -0.217 | -0.215 | -0.214 | -1.286 | 0.215 | -1.527 | 0.144 | -0.41 | 0.686 |
| **MMP.1** | -0.092 | -0.089 | -0.093 | -2.318 | 0.032 | 0.898 | 0.381 | 2.771 | 0.013 |
| **MRC2** | 0.044 | 0.041 | 0.044 | 2.357 | 0.03 | -0.422 | 0.678 | -2.275 | 0.035 |
| **Myeloperoxi dase** | 0 | 0 | 0.001 | 0.37 | 0.715 | -0.551 | 0.589 | -0.925 | 0.367 |
| **N.CDase** | 0.081 | 0.082 | 0.081 | -1.712 | 0.104 | -0.046 | 0.964 | 0.88 | 0.39 |
| **NCAM.120** | 0.05 | 0.05 | 0.049 | 0.1 | 0.922 | 0.356 | 0.726 | 0.41 | 0.687 |
| **NEP** | -0.021 | -0.024 | -0.022 | 2.022 | 0.058 | 0.482 | 0.636 | -0.937 | 0.361 |
| **NMNAT1** | -0.049 | -0.049 | -0.05 | -0.979 | 0.341 | 1.429 | 0.17 | 2.358 | 0.03 |
| **NOTCH1** | 0.068 | 0.066 | 0.068 | 3.102 | 0.006 | 0.666 | 0.514 | -1.976 | 0.064 |
| **NRTK3** | 0.148 | 0.149 | 0.149 | -0.641 | 0.529 | -0.675 | 0.508 | 0.123 | 0.903 |
| **NTRK3** | 0.148 | 0.146 | 0.146 | 1.672 | 0.112 | 2.026 | 0.058 | 0.072 | 0.943 |
| **OSM** | 0.092 | 0.096 | 0.089 | -2.033 | 0.057 | 1.04 | 0.312 | 3.606 | 0.002 |

| | | | | | | | | | |
|---|---|---|---|---|---|---|---|---|---|
| Osteomodulin | 0.337 | 0.337 | 0.335 | 0.295 | 0.771 | 1.524 | 0.145 | 1.095 | 0.288 |
| PAPP.A | -0.423 | -0.418 | -0.423 | -2.139 | 0.046 | -0.145 | 0.887 | 2.366 | 0.029 |
| PIGR | -0.23 | -0.227 | -0.225 | -1.709 | 0.105 | -3.536 | 0.002 | -1.347 | 0.195 |
| RARRES2 | 0.009 | 0.009 | 0.012 | 0.193 | 0.849 | -1.808 | 0.087 | -1.999 | 0.061 |
| Resistin | -0.122 | -0.116 | -0.123 | -2.906 | 0.009 | 0.487 | 0.632 | 3.396 | 0.003 |
| S100.A9 | 0.028 | 0.03 | 0.029 | -0.762 | 0.456 | -0.309 | 0.761 | 0.485 | 0.634 |
| Semaphorin.3E | -0.051 | -0.054 | -0.053 | 3.041 | 0.007 | 2.608 | 0.018 | -0.98 | 0.34 |
| SERPIN.A3 | 0.154 | 0.151 | 0.152 | 1.699 | 0.107 | 0.927 | 0.366 | -0.497 | 0.625 |
| SHBG | -0.059 | -0.057 | -0.06 | -2.044 | 0.056 | 0.478 | 0.639 | 2.337 | 0.031 |
| SIGLEC1 | -0.042 | -0.037 | -0.043 | -3.971 | 0.001 | 1.001 | 0.33 | 4.408 | 0.00034 |
| SKR3 | 0.155 | 0.153 | 0.157 | 1.163 | 0.26 | -2.452 | 0.025 | -3.331 | 0.004 |
| SLITRK5 | 0.162 | 0.157 | 0.161 | 3.63 | 0.002 | 0.986 | 0.337 | -2.323 | 0.032 |
| SMPD1 | -0.032 | -0.03 | -0.032 | -1.815 | 0.086 | -0.376 | 0.711 | 1.419 | 0.173 |
| Stanniocalcin.1 | -0.046 | -0.047 | -0.044 | 1.562 | 0.136 | -2.293 | 0.034 | -4.345 | 0.00039 |
| Testican.2 | -0.212 | -0.212 | -0.211 | 0.463 | 0.649 | -0.394 | 0.698 | -1.099 | 0.286 |
| TGF.alpha | 0.021 | 0.026 | 0.018 | -2.842 | 0.011 | 1.29 | 0.213 | 3.989 | 0.001 |
| THBS2 | -0.123 | -0.125 | -0.121 | 1.135 | 0.271 | -2.185 | 0.042 | -2.637 | 0.017 |
| TNFRSF17 | 0.017 | 0.018 | 0.017 | -1.363 | 0.19 | -0.288 | 0.776 | 1.74 | 0.099 |
| TNFRSF1B | -0.106 | -0.104 | -0.105 | -1.898 | 0.074 | -1.028 | 0.317 | 0.525 | 0.606 |
| TPO | -0.309 | -0.306 | -0.306 | -2.217 | 0.04 | -2.607 | 0.018 | -0.236 | 0.816 |
| Trypsin.2 | -0.104 | -0.103 | -0.104 | -0.609 | 0.55 | 0.494 | 0.627 | 0.859 | 0.402 |
| Tryptase.beta.2 | -0.218 | -0.222 | -0.214 | 2.005 | 0.06 | -1.544 | 0.14 | -3.893 | 0.001 |
| VCAM1 | -0.01 | -0.011 | -0.01 | 2.165 | 0.044 | 0.595 | 0.559 | -1.002 | 0.33 |
| VEGFA | 0.128 | 0.131 | 0.128 | -2.187 | 0.042 | 0.076 | 0.941 | 2.474 | 0.024 |
| WFIKKN2 | 0.051 | 0.05 | 0.049 | 0.993 | 0.334 | 2.972 | 0.008 | 1.888 | 0.075 |
| Relative.IL6.Level | -0.079 | -0.086 | -0.041 | 0.501 | 0.622 | -2.495 | 0.023 | -3.292 | 0.004 |

**Table S3. Statistical analysis for comparing baseline and 6-month Marioni marker proportions in the Dasatinib, Quercetin, and Fisetin Study.** The first two columns show the mean values for each immune cell proportion at each time point. The next columns have information about the t-test between baseline and 6-month test.

| | Mean | | Baseline vs 6-month | |
|---|---|---|---|---|
| | Baseline | 6-month | T-score | P-value |
| Epigenetic Age Zhang. | -0.679 | -0.556 | -0.719 | 0.492 |
| Alcohol | -11.613 | -11.406 | -2.928 | 0.019 |
| Body Fat | -8.756 | -8.987 | 0.247 | 0.811 |
| Body Mass Index | -0.571 | -0.58 | 0.36 | 0.728 |
| HDL Cholesterol | 2.598 | 2.612 | -0.313 | 0.762 |

| | | | | |
|---|---|---|---|---|
| **Smoking** | 2.699 | 2.581 | 1.45 | 0.185 |
| **Waist.Hip.Ratio** | -0.323 | -0.306 | -1.891 | 0.095 |
| **ADAMTS** | 0.1 | 0.104 | -1.311 | 0.226 |
| **Adiponectin** | -0.072 | -0.073 | 0.49 | 0.637 |
| **Afamin** | -0.01 | -0.01 | -0.279 | 0.787 |
| **Alpha L iduronidase** | 0.112 | 0.112 | -0.082 | 0.937 |
| **Aminoacylase 1** | -0.319 | -0.322 | 0.687 | 0.511 |
| **B2 microglobulin** | -0.376 | -0.376 | 0.066 | 0.949 |
| **BMP.1** | 0.115 | 0.121 | -2.644 | 0.03 |
| **CCL11** | -0.003 | -0.004 | 0.362 | 0.726 |
| **CCL17** | -0.428 | -0.431 | 1.266 | 0.241 |
| **CCL18** | -0.145 | -0.144 | -0.892 | 0.399 |
| **CCL21** | -0.131 | -0.133 | 0.968 | 0.362 |
| **CCL22** | -0.065 | -0.066 | 0.337 | 0.745 |
| **CCL25.C.C** | -0.061 | -0.062 | 1.089 | 0.308 |
| **CD163** | -0.246 | -0.244 | -0.666 | 0.524 |
| **CD209.antigen** | 0.134 | 0.139 | -1.453 | 0.184 |
| **CD48.antigen** | -0.157 | -0.158 | 0.267 | 0.797 |
| **CD6** | 0.097 | 0.096 | 0.342 | 0.741 |
| **CDL5** | -0.069 | -0.07 | 0.234 | 0.821 |
| **CHIT.1** | -0.173 | -0.173 | -0.071 | 0.945 |
| **CLEC11A.e1** | -0.005 | -0.015 | 1.792 | 0.111 |
| **CLEC11A.e2** | -0.145 | -0.153 | 1.798 | 0.11 |
| **Coagulation.factor.VII** | 0.049 | 0.048 | 0.353 | 0.733 |
| **Complement.C4** | 0.036 | 0.037 | -1.052 | 0.324 |
| **Complement.C5a** | 0.159 | 0.161 | -0.498 | 0.632 |
| **Complement.c9** | -0.009 | -0.013 | 0.756 | 0.471 |
| **Contactin.4** | 0.171 | 0.17 | 0.386 | 0.71 |
| **CRP** | -0.113 | -0.117 | 1.396 | 0.2 |
| **CRTAM** | 0.053 | 0.055 | -0.797 | 0.448 |
| **CXCL10** | 0.137 | 0.141 | -0.95 | 0.37 |
| **CXCL10.soma** | -0.347 | -0.346 | -0.206 | 0.842 |
| **CXCL11** | 0.097 | 0.101 | -0.703 | 0.502 |
| **CXCL11.soma** | -0.056 | -0.054 | -0.788 | 0.453 |
| **CXCL9** | -0.037 | -0.035 | -1.532 | 0.164 |
| **E.selectin** | -0.026 | -0.026 | -0.018 | 0.986 |
| **Ectodysplasin.A** | -0.216 | -0.219 | 1.531 | 0.164 |
| **EN.RAGE** | 0.051 | 0.047 | 1.278 | 0.237 |
| **ENPP7** | -0.017 | -0.015 | -0.203 | 0.844 |
| **ESM.1** | -0.244 | -0.247 | 1.814 | 0.107 |

| | | | | |
|---|---|---|---|---|
| **EZR** | -0.019 | -0.018 | -0.495 | 0.634 |
| **FAP** | -0.143 | -0.141 | -1.418 | 0.194 |
| **FCER2** | -0.251 | -0.252 | 0.271 | 0.793 |
| **FCGR3A** | -0.203 | -0.207 | 1.06 | 0.32 |
| **FcRL2** | -0.175 | -0.176 | 0.479 | 0.645 |
| **FGF.21** | -0.116 | -0.113 | -1.642 | 0.139 |
| **G.CSF** | -0.015 | -0.016 | 0.418 | 0.687 |
| **Galectin.4** | -0.208 | -0.208 | -0.302 | 0.771 |
| **GDF.8** | 0.141 | 0.142 | -0.852 | 0.419 |
| **GHR** | 0.139 | 0.141 | -0.643 | 0.538 |
| **GPIba** | -0.362 | -0.363 | 0.141 | 0.892 |
| **Granulysin** | -0.115 | -0.112 | -0.76 | 0.469 |
| **Granzyme.A** | -0.034 | -0.035 | 0.326 | 0.753 |
| **GZMA** | -0.067 | -0.068 | 0.762 | 0.468 |
| **HCII** | 0.039 | 0.038 | 0.084 | 0.935 |
| **HGF** | 0.043 | 0.041 | 0.824 | 0.434 |
| **HGFA** | 0.358 | 0.359 | -0.499 | 0.631 |
| **HGFI** | 0.551 | 0.56 | -0.88 | 0.405 |
| **ICAM5** | -0.092 | -0.093 | 0.203 | 0.844 |
| **IGFBP.1** | -0.124 | -0.125 | 0.283 | 0.784 |
| **IGFBP.4** | -0.048 | -0.045 | -3.234 | 0.012 |
| **Insulin.receptor** | -0.136 | -0.137 | 0.54 | 0.604 |
| **Interleukin.19** | -0.011 | -0.009 | -0.67 | 0.522 |
| **L.selectin** | 0.051 | 0.051 | -0.206 | 0.842 |
| **LFT** | -0.005 | -0.007 | 0.747 | 0.476 |
| **LGALS3BP** | 0.008 | 0.006 | 1.019 | 0.338 |
| **LY9** | -0.124 | -0.123 | -0.265 | 0.798 |
| **Lymphotoxin.abeta** | -0.037 | -0.034 | -0.991 | 0.351 |
| **MIA** | 0.102 | 0.103 | -0.456 | 0.66 |
| **MMP.1.1** | -0.123 | -0.122 | -0.348 | 0.737 |
| **MMP.12** | -0.201 | -0.201 | 0.01 | 0.992 |
| **MMP.9** | -0.211 | -0.212 | 0.323 | 0.755 |
| **MMP.1** | -0.097 | -0.098 | 0.689 | 0.51 |
| **MRC2** | 0.043 | 0.043 | 0.011 | 0.992 |
| **Myeloperoxidase** | -0.001 | -0.001 | -0.047 | 0.964 |
| **N.CDase** | 0.105 | 0.106 | -0.572 | 0.583 |
| **NCAM.120** | 0.043 | 0.043 | 0.294 | 0.776 |
| **NEP** | -0.037 | -0.031 | -2.053 | 0.074 |
| **NMNAT1** | -0.05 | -0.05 | -0.392 | 0.705 |
| **NOTCH1** | 0.069 | 0.07 | -0.492 | 0.636 |

| | | | | |
|---|---|---|---|---|
| **NRTK3** | 0.144 | 0.147 | -1.144 | 0.286 |
| **NTRK3** | 0.136 | 0.135 | 0.234 | 0.821 |
| **OSM** | 0.1 | 0.094 | 1.746 | 0.119 |
| **Osteomodulin** | 0.329 | 0.326 | 1.924 | 0.091 |
| **PAPP.A** | -0.402 | -0.399 | -0.426 | 0.681 |
| **PIGR** | -0.216 | -0.218 | 0.636 | 0.542 |
| **RARRES2** | 0.01 | 0.013 | -1.15 | 0.283 |
| **Resistin** | -0.122 | -0.126 | 0.93 | 0.379 |
| **S100.A9** | 0.035 | 0.034 | 0.345 | 0.739 |
| **Semaphorin.3E** | -0.057 | -0.057 | 0.245 | 0.813 |
| **SERPIN.A3** | 0.157 | 0.157 | 0.156 | 0.88 |
| **SHBG** | -0.068 | -0.07 | 0.834 | 0.428 |
| **SIGLEC1** | -0.043 | -0.042 | -0.524 | 0.614 |
| **SKR3** | 0.163 | 0.164 | -0.485 | 0.641 |
| **SLITRK5** | 0.148 | 0.154 | -1.471 | 0.18 |
| **SMPD1** | -0.034 | -0.034 | 0.06 | 0.953 |
| **Stanniocalcin.1** | -0.04 | -0.037 | -1.222 | 0.257 |
| **Testican.2** | -0.212 | -0.214 | 0.795 | 0.449 |
| **TGF.alpha** | 0.024 | 0.019 | 1.798 | 0.11 |
| **THBS2** | -0.116 | -0.119 | 0.996 | 0.348 |
| **TNFRSF17** | 0.013 | 0.012 | 1.069 | 0.316 |
| **TNFRSF1B** | -0.106 | -0.108 | 1.039 | 0.329 |
| **TPO** | -0.308 | -0.31 | 0.7 | 0.504 |
| **Trypsin.2** | -0.104 | -0.103 | -1.352 | 0.213 |
| **Tryptase.beta.2** | -0.174 | -0.174 | 0.043 | 0.967 |
| **VCAM1** | -0.008 | -0.008 | 0.012 | 0.991 |
| **VEGFA** | 0.125 | 0.124 | 0.255 | 0.805 |
