## Supplementary material for "Exploring the effects of Dasatinib, Quercetin, and Fisetin on DNA methylation clocks: a longitudinal study on senolytic interventions": Tables

**Table 1. Characteristics of participants in both trials.**

| | Dasatinib and Quercetin study | Dasatinib, Quercetin, and Fisetin study |
|---|---|---|
| **Sample Size** | 19 | 19 |
| **Age in years, mean (range)** | 59.6 (43.0 - 86.6) | 60.9 (44.5 - 88.0) |
| **Sex, male** | 11 (57.9%) | 8 (42.1%) |

**Table 2. Statistical analysis for comparing baseline, 3 months, and 6 months epigenetic age acceleration (EAA) in the Dasatinib and Quercetin Study.** The first three columns show the mean values for each EAA clock at each time point. The next columns have information about the t-test between baseline and 3-month test, between baseline and 6-month test, and between 3-month and 6-month tests, respectively.

| | Mean | | | Baseline vs 3-month | | Baseline vs 6-month | | 3-month vs 6-month | |
|---|---|---|---|---|---|---|---|---|---|
| | **Base** | **3m** | **6m** | **T-score** | **P-value** | **T-score** | **P-value** | **T-score** | **P-value** |
| **PC Horvath pan tissue EAA** | -1.485 | 1.254 | -0.926 | -6.258 | $6.7 \cdot 10^{-6}$ | -1.230 | 0.234 | 4.527 | $2.6 \cdot 10^{-4}$ |
| **PC Horvath Skin and Blood EAA** | -0.625 | 0.194 | -0.064 | -2.450 | 0.025 | -1.075 | 0.297 | 0.598 | 0.557 |
| **PC Hannum EAA** | -1.165 | 0.474 | 0.184 | -3.622 | 0.002 | -2.561 | 0.020 | 0.546 | 0.592 |
| **PC GrimAge EAA** | -0.173 | -0.179 | 0.374 | 0.016 | 0.988 | -1.431 | 0.170 | -1.476 | 0.157 |
| **PC DNAmPheno Age EAA** | -1.766 | 0.891 | 0.134 | -3.237 | 0.005 | -2.240 | 0.038 | 0.936 | 0.362 |
| **PC DNAmTL EAA** | 0.043 | -0.024 | -0.010 | 5.098 | $7.5 \cdot 10^{-5}$ | 4.286 | $4.4 \cdot 10^{-4}$ | -0.931 | 0.364 |
| **DunedinPACE** | 0.929 | 0.923 | 0.927 | 0.392 | 0.699 | 0.112 | 0.912 | -0.230 | 0.821 |

| | | | | | | | | | |
|---|---|---|---|---|---|---|---|---|---|
| **Intrinclock EAA** | 0.039 | -0.428 | 0.389 | 0.714 | 0.484 | -0.499 | 0.624 | -1.235 | 0.233 |

**Table 3. Statistical analysis for comparing baseline and 6-month epigenetic age acceleration (EAA) in the Dasatinib, Quercetin, and Fisetin Study.** The first two columns show the mean values for each EAA clock at each time point. The next columns have information about the t-test between baseline and 6-month test.

| | **Mean** | | **Baseline vs 6-month** | |
|---|---|---|---|---|
| | **Baseline** | **6-month** | **T-score** | **P-value** |
| **PC Horvath pan tissue EAA** | -0.201 | 1.607 | -2.643 | 0.017 |
| **PC Horvath Skin and Blood EAA** | -0.057 | 0.826 | -1.838 | 0.083 |
| **PC Hannum EAA** | -0.162 | 0.853 | -1.774 | 0.093 |
| **PC GrimAge EAA** | 0.101 | 0.057 | 0.140 | 0.890 |
| **PC DNAmPhenoAge EAA** | -0.033 | 0.964 | -1.094 | 0.288 |
| **PC DNAmTL EAA** | 0.005 | -0.027 | 1.698 | 0.107 |
| **DunedinPACE** | 0.937 | 0.918 | 1.016 | 0.323 |
| **Intrinclock EAA** | 0.637 | -0.637 | 1.608 | 0.125 |

**Table 4. Statistical analysis for comparing baseline, 3 months, and 6 months immune cell proportions in the Dasatinib and Quercetin Study.** The first three columns show the mean values for each immune cell proportion at each time point. The next columns have information about the t-test between baseline and 3-month test, between baseline and 6-month test, and between 3-month and 6-month tests, respectively.

| | **Mean** | | | **Baseline vs 3-month** | | **Baseline vs 6-month** | | **3-month vs 6-month** | |
|---|---|---|---|---|---|---|---|---|---|
| | **Base** | **3m** | **6m** | **T-score** | **P-value** | **T-score** | **P-value** | **T-score** | **P-value** |
| **CD4T naive cells** | 0.077 | 0.074 | 0.060 | 0.493 | 0.628 | 2.228 | 0.039 | 2.364 | 0.029 |

| | | | | | | | | | |
|---|---|---|---|---|---|---|---|---|---|
| **Basophiles** | 0.020 | 0.021 | 0.020 | -1.147 | 0.266 | -0.408 | 0.688 | 0.439 | 0.666 |
| **CD4T memory cells** | 0.084 | 0.081 | 0.079 | 0.535 | 0.599 | 0.639 | 0.531 | 0.410 | 0.686 |
| **B memory cells** | 0.019 | 0.017 | 0.019 | 1.287 | 0.214 | -0.457 | 0.653 | -1.380 | 0.184 |
| **B naive cells** | 0.040 | 0.035 | 0.046 | 1.611 | 0.125 | -2.018 | 0.059 | -3.907 | 0.001 |
| **T regulatory cells** | 0.005 | 0.007 | 0.008 | -0.978 | 0.341 | -1.946 | 0.067 | -0.842 | 0.411 |
| **CD8T memory cells** | 0.051 | 0.052 | 0.049 | -0.290 | 0.775 | 0.359 | 0.724 | 0.650 | 0.524 |
| **CD8T naive cells** | 0.025 | 0.028 | 0.026 | -1.354 | 0.193 | -0.456 | 0.654 | 0.607 | 0.552 |
| **Eosinophiles** | 0.009 | 0.009 | 0.005 | 0.306 | 0.763 | 1.400 | 0.178 | 1.036 | 0.314 |
| **Natural Killer** | 0.044 | 0.047 | 0.048 | -0.696 | 0.495 | -1.046 | 0.309 | -0.339 | 0.739 |
| **Neutrophiles** | 0.564 | 0.575 | 0.556 | -0.612 | 0.548 | 0.309 | 0.761 | 0.966 | 0.347 |
| **Monocytes** | 0.062 | 0.053 | 0.082 | 2.282 | 0.035 | -3.415 | 0.003 | -5.527 | $3.0\cdot10^{-5}$ |

**Table 5. Statistical analysis for comparing baseline and 6-month immune cell proportions in the Dasatinib, Quercetin, and Fisetin Study.** The first two columns show the mean values for each immune cell proportion at each time point. The next columns have information about the t-test between baseline and 6-month test.

| | Mean | | Baseline vs 6-month | |
|---|---|---|---|---|
| | **Baseline** | **6-month** | **T-score** | **P-value** |
| **CD4T naive cells** | 0.065 | 0.063 | 0.200 | 0.844 |
| **Basophiles** | 0.014 | 0.013 | 0.507 | 0.618 |
| **CD4T memory cells** | 0.081 | 0.089 | -0.926 | 0.367 |
| **B memory cells** | 0.017 | 0.015 | 1.122 | 0.277 |
| **B naive cells** | 0.040 | 0.024 | 4.470 | $3.0\cdot10^{-4}$ |

| | | | | |
|---|---|---|---|---|
| **T regulatory cells** | 0.006 | 0.004 | 2.069 | 0.053 |
| **CD8T memory cells** | 0.063 | 0.058 | 0.950 | 0.354 |
| **CD8T naive cells** | 0.020 | 0.017 | 1.202 | 0.245 |
| **Eosinophiles** | 0.011 | 0.011 | -0.009 | 0.993 |
| **Natural Killer** | 0.055 | 0.048 | 1.620 | 0.123 |
| **Neutrophiles** | 0.569 | 0.597 | -1.336 | 0.198 |
| **Monocytes** | 0.060 | 0.062 | -0.421 | 0.679 |

**Table 6. Inflammation and inflammatory proteomic EpiScore analysis between baseline, 3-month test, and 6-month test in the Dasatinib and Quercetin (DQ) trial.** The first three columns show the mean values for each protein methylation risk score (MRS) at each time point. The next columns have information about the t-test between baseline and 3-month test, between baseline and 6-month test, and between 3-month and 6-month tests, respectively.

| | **Mean** | | | **Baseline vs 3-month** | | **Baseline vs 6-month** | | **3-month vs 6-month** | |
|---|---|---|---|---|---|---|---|---|---|
| | **Base** | **3m** | **6m** | **T-score** | **P-value** | **T-score** | **P-value** | **T-score** | **P-value** |
| **CCL11** | -0.007 | -0.007 | -0.004 | 0.071 | 0.944 | -2.94 | 0.009 | -3.97 | 0.001 |
| **CCL17** | -0.434 | -0.428 | -0.435 | -3.306 | 0.004 | 1.062 | 0.302 | 3.614 | 0.002 |
| **CCL18** | -0.15 | -0.15 | -0.148 | 0.002 | 0.998 | -1.443 | 0.166 | -1.722 | 0.102 |
| **CCL21** | -0.127 | -0.128 | -0.127 | 1.449 | 0.165 | 0.06 | 0.953 | -1.473 | 0.158 |
| **CCL22** | -0.066 | -0.065 | -0.064 | -0.428 | 0.674 | -3.241 | 0.005 | -3.269 | 0.004 |
| **Complement C4** | 0.032 | 0.031 | 0.033 | 2.923 | 0.009 | -1.723 | 0.102 | -4.513 | $2.7\cdot10^{-4}$ |
| **Complement C5a** | 0.155 | 0.148 | 0.158 | 3.721 | 0.002 | -2.268 | 0.036 | -6.877 | $2.0\cdot10^{-5}$ |
| **Complement C9** | -0.013 | -0.012 | -0.011 | -0.545 | 0.592 | -0.751 | 0.462 | -0.229 | 0.822 |
| **CRP** | -0.114 | -0.107 | -0.115 | -4.888 | $1.2\cdot10^{-4}$ | 0.979 | 0.34 | 4.679 | $1.9\cdot10^{-4}$ |

| | | | | | | | | | |
|---|---|---|---|---|---|---|---|---|---|
| **CXCL10 soma** | -0.345 | -0.338 | -0.343 | -2.939 | 0.009 | -0.966 | 0.347 | 1.911 | 0.072 |
| **CXCL11 soma** | -0.053 | -0.05 | -0.054 | -3.291 | 0.004 | 1.282 | 0.216 | 5.23 | $5.7 \cdot 10^{-5}$ |
| **CXCL9** | -0.034 | -0.031 | -0.037 | -3.347 | 0.004 | 2.728 | 0.014 | 5.132 | $7.0 \cdot 10^{-5}$ |
| **Interleukin 19** | -0.002 | -0.005 | -0.005 | 1.064 | 0.302 | 1.241 | 0.231 | 0.188 | 0.853 |
| **TGF alpha** | 0.021 | 0.026 | 0.018 | -2.842 | 0.011 | 1.29 | 0.213 | 3.989 | 0.001 |
| **TNFRSF1B** | -0.106 | -0.104 | -0.105 | -1.898 | 0.074 | -1.028 | 0.317 | 0.525 | 0.606 |
| **Relative IL6 Level** | -0.079 | -0.086 | -0.041 | 0.501 | 0.622 | -2.495 | 0.023 | -3.292 | 0.004 |
